# Cost-effectiveness of Stockholm3 test and magnetic resonance imaging in prostate cancer screening: a microsimulation study

**DOI:** 10.1101/2021.03.31.21254617

**Authors:** Shuang Hao, Emelie Heintz, Ellinor Östensson, Andrea Discacciati, Fredrik Jäderling, Henrik Grönberg, Martin Eklund, Tobias Nordström, Mark Clements

## Abstract

**Objective:** Assess the cost-effectiveness of no screening and quadrennial magnetic resonance imaging (MRI)-based screening for prostate cancer using either Stockholm3 or prostate-specific antigen (PSA) test as a reflex test.

**Methods:** Test characteristics were estimated from the STHLM3-MR study (NCT03377881). A cost-utility analysis was conducted from a lifetime societal perspective using a microsimulation model for men aged 55-69 in Sweden for no screening and three quadrennial screening strategies, including: PSA≥3ng/mL; and Stockholm3 with reflex test thresholds of PSA≥1.5 and 2ng/mL. Men with a positive test had an MRI, and those MRI positive had combined targeted and systematic biopsies. Predictions included the number of tests, cancer incidence and mortality, costs, quality-adjusted life-years (QALYs) and incremental cost-effectiveness ratios (ICERs). Uncertainties in key parameters were assessed using sensitivity analyses.

**Results:** Compared with no screening, the screening strategies were predicted to reduce prostate cancer deaths by 7-9% across a lifetime and were considered to be moderate costs per QALY gained in Sweden. Using Stockholm3 with a reflex threshold of PSA≥2ng/mL resulted in a 60% reduction in MRI compared with screening using PSA. This Stockholm3 strategy was cost-effective with a probability of 70% at a cost-effectiveness threshold of €47,218 (500,000 SEK).

**Conclusions:** All screening strategies were considered to be moderate costs per QALY gained compared with no screening. Screening with Stockholm3 test at a reflex threshold of PSA≥2ng/mL and MRI was predicted to be cost-effective in Sweden. Use of the Stockholm3 test may reduce screening-related harms and costs while maintaining the health benefits from early detection.

## Background

Prostate cancer (PCa) is the most common male cancer diagnosed and the leading cause of cancer death in Sweden ^1^. Organised screening for prostate cancer with the prostate-specific antigen (PSA) test remains controversial. A 20% mortality reduction from PSA screening was found by the European Randomized Study of Screening for Prostate Cancer (ERSPC) compared with no screening after 16-year follow up ^2^. However, 39% of the men diagnosed in the ERSPC had low risk cancers, with either clinical T1 stage or International Society of Urological Pathology (ISUP) Grade Group 1 (GG=1) histopathology ^2^. Apart from the over-diagnosis of low-risk cancers, other potential risks of PSA screening include unnecessary biopsies and over-treatment, which are accompanied by a reduction in health-related quality of life and an increase in the economic burden ^3,4^. Given these uncertainties, a number of clinical guidelines have recommended shared decision-making for whether to start PCa testing.

The availability of new diagnostic tests offers a potential of risk-stratified screening for PCa in Sweden. The Stockholm3 (S3M) test is a risk-model that combines PSA, single nucleotide polymorphisms, clinical variables, as well as established and novel plasma protein biomarkers ^5^. The STHLM3 diagnostic study (ISRCTN84445406) compared Stockholm3 with PSA testing within the context of systematic biopsies. Compared to men with PSA 3-10 ng/mL, the Stockholm3 test with a reflex threshold of PSA≥1 ng/mL maintained the sensitivity to detect ISUP GG≥2 cancers, and reduced benign biopsies by 44% and biopsies in detecting ISUP GG=1 cancers by 17% ^5^. Compared to men with PSA 3-10ng/mL, the Stockholm3 test with a reflex threshold of PSA≥2ng/mL had fewer biopsies with reductions of 52%, 28% and 5% in detecting benign biopsies (represented GG=0), GG=1 and GG≥2 cancers, respectively ^6^.

The use of magnetic resonance imaging (MRI) to complement PCa diagnosis has been recommended by Swedish guidelines ^7^ and the European Association of Urology (EAU) ^8^ due to its improved sensitivity and specificity ^9,10^. For biopsy-naïve patients, the combined MRI-guided targeted biopsy (TBx) and systematic biopsy (SBx) was recommended by EAU for those who have Prostate Imaging Reporting and Data System (PI-RADS) scores 3-5 of MRI ^8^. The performance of using either a Stockholm3 or PSA test with MRI for PCa diagnosis has been examined by the recent STHLM3-MR study (NCT03377881). Men with S3M≥11% or PSA≥3ng/mL were randomised to have either SBx in the standard arm or MRI with a combined TBx/SBx in the experimental arm. PSA≥1.5ng/mL was used as the reflex threshold for Stockholm3 test. Men with S3M≥15% and a positive MRI result (MRI+) were referred to undertake TBx/SBx and those with a negative MRI result (MRI-) but a high value of S3M≥25% were referred for a SBx. Compared with PSA≥3 and using an intention-to-treat (ITT) perspective, using PSA≥1.5ng/mL and S3M≥15% had the same sensitivity in detecting ISUP GG≥2 cancers, while ISUP GG=1 cancers and benign biopsies were reduced by 17% and 19%, respectively (Nordström et al, submitted, 2021) ^11^.

From a cost-effectiveness perspective, given quadrennial PCa screening for men aged 55-69 with systematic biopsies in Sweden, we predicted that Stockholm3 with a reflex threshold of PSA 2ng/mL would have an incremental cost-effectiveness ratio (ICER) of €6,000 per quality-adjusted life year (QALY) gained compared with PSA screening ^6^. Moreover, for PSA screening with a threshold of PSA≥3ng/mL, we predicted that PSA+MRI+TBx/SBx would be strongly dominant over PSA+SBx with higher QALY gains and lower costs across a lifetime (Hao et al, under review, 2021). Employing MRI+TBx/SBx in addition to Stockholm3 for screening is expected to further reduce unnecessary biopsies and maintain effectiveness.

The National Board of Health and Welfare in Sweden has established a national pilot project of organised prostate cancer testing with MRI ^12^. To support national policy decision-making on organised population-based screening for PCa, this study aimed to assess the cost-effectiveness of Stockholm3 compared with PSA, given quadrennial screening, MRI and combined TBx/SBx among men aged 55 to 69 years in Sweden.

## Methods

### Study population and screening strategies

Following the primary screening protocol of ERSPC, quadrennial screening strategies for men aged 55-69 years were assumed to be administered by general practitioners with referral to a radiologist for an MRI, followed by a referral to an urologist for further decision on biopsy, based on the PI-RADS score and Stockholm3 results. The four strategies (Figure 1) were: I) No screening (symptomatic cancer detection only); II) MRI for PSA≥3ng/mL (PSA+MRI+TBx/SBx) and TBx/SBx for PI-RADS 3-5; III) MRI for S3M≥15% using a reflex threshold of PSA≥1.5ng/mL and TBx/SBx for men who had PI-RADS 3-5 (PSA1.5+S3M+MRI+TBx/SBx); and IV) MRI for S3M≥15% using a reflex threshold of PSA≥2ng/mL and TBx/SBx for men who had PI-RADS 3-5 (PSA2+S3M+MRI+TBx/SBx). Strategy I was used as a comparator with the screening strategies.

**Figure 1:**
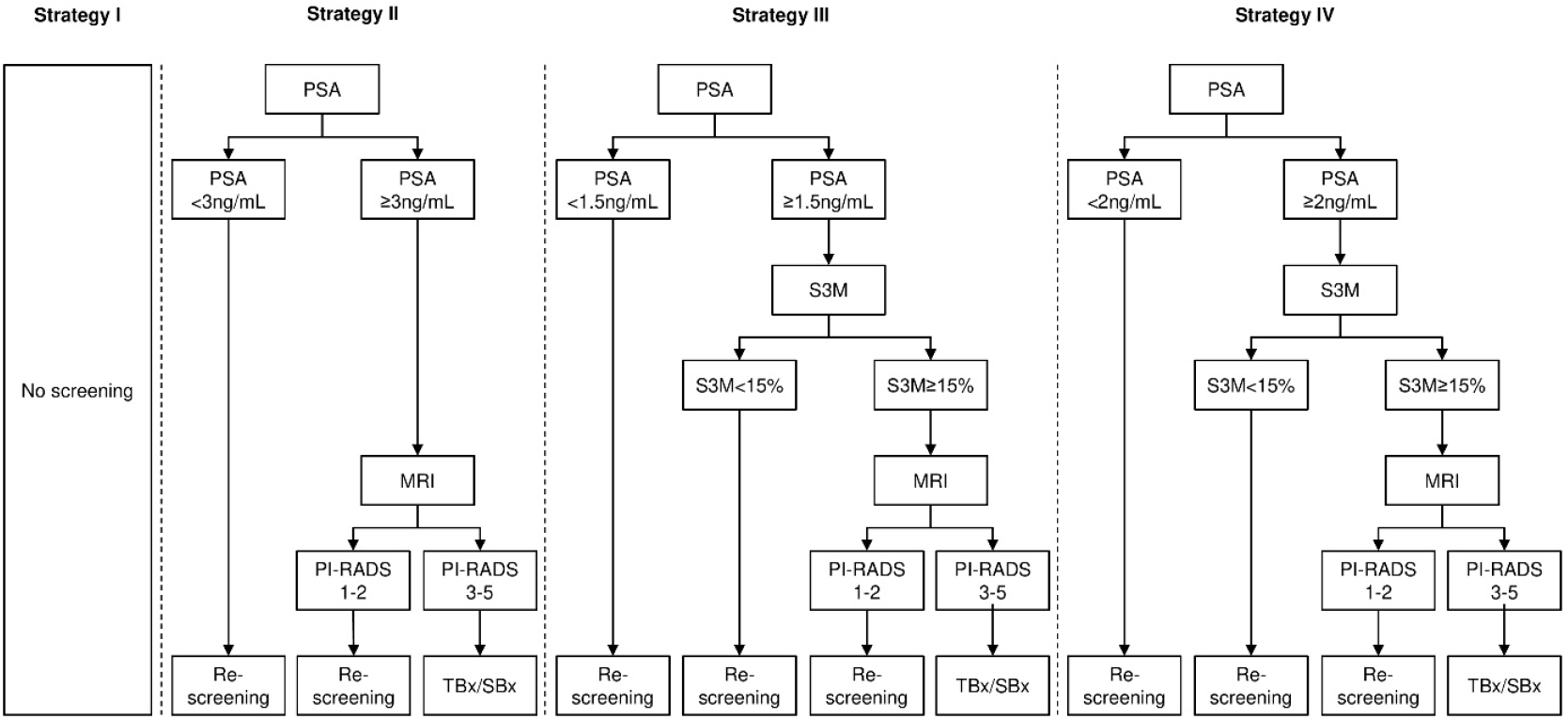
Illustrations of strategies. Legend: Strategy I: no screening; Strategy II: quadrennial screening using MRI on PSA≥3ng/mL, with combined TBx/SBx on positive MRI results; Strategy III: quadrennial screening using S3M on PSA≥1.5ng/ml and MRI on S3M≥15%, with TBx/SBx on positive MRI results; Strategy IV: quadrennial screening using S3M on PSA≥2ng/mL and MRI on S3M≥15%, with TBx/SBx on positive MRI results; MRI: Magnetic Resonance Imaging; PI-RADS: Prostate Imaging Reporting and Data System; PSA: Prostate-Specific Antigen; SBx: Systematic Biopsy; S3M: Stockholm3 test; TBx: Targeted Biopsy; TBx/SBx: combined Targeted and Systematic Biopsies

### Simulation model

We used an open source microsimulation model to simulate the life histories of men with PCa, including PSA testing, disease onset, progression, diagnosis, treatment and death (see Figure S1, Supplementary Material I) ^13^. Model inputs include biopsy and treatment pathways informed by data from the Stockholm PSA and Biopsy Register ^14^. The natural history model was calibrated to prostate cancer incidence in Sweden and the effect of screening and biopsy compliance from ERSPC. Men in a preclinical state were assumed to be asymptomatic and their states were defined by the ISUP grade group (GG1, GG2-3, GG4-5), T-stage (T1-T2, T3-T4) and metastasis. The cancer may progress between preclinical states (although the model does not allow for grade de-differentiation) and be detected clinically. Treatment pathways in the clinical stage included active surveillance, radical prostatectomy, radiation therapy, post-treatment follow-up, metastatic drug treatment, palliative therapy and terminal care (see Supplementary Material II for a diagram of the screening and treatment pathways). Detailed description of the model can be found by Karlsson et al ^13^. This model provides internally consistent contrasts between interventions.

### Test Characteristics

Test characteristics were estimated based on data from the experimental arm of STHLM3-MR study that matched our strategies II to IV. Per the protocol of STHLM3-MR study, men with an MRI without visible lesions (PI-RADS≤2) but a high risk of prostate cancer (S3M≥25%) were referred for systematic biopsies ^11^. Our baseline analysis used a per-protocol (PP) perspective and excluded the biopsy findings for men with a negative MRI result and S3M≥25%. The test characteristics for the protocol that includes negative MRI results and S3M≥25% were shown in the Supplementary Material Table S2. Patients with positive MRI results and ISUP grading from both TBx and SBx were included for the calculation of detected cases of ISUP GG=0, GG=1 and GG≥2 cancers. Patients who undertook MRI with negative results did not have a biopsy. For the relative positive fractions comparing the screening strategies, see Supplementary Material I Table S3.

We also estimated the probability of positive MRI result (MRI+) given: (i) a positive PSA result and GG; (ii) a reflex threshold of Stockholm3 test as PSA≥1.5 ng/mL and GG and (iii) a reflex threshold of Stockholm3 test as PSA≥2 ng/mL and GG. Note that GG was defined using the maximum grading from TBx and SBx. Due to the study design where men with PI-RADS 1-2 did not undertake SBx, there a was lack of information on the true negative of GG=0 and true positive of GG=1 and GG≥2 cancers on negative MRI results. Therefore, the total number of benign biopsies (GG=0) might be overestimated and the total number of GG=1 and GG≥2 cancers might be underestimated when using TBx/SBx on MRI positive results only. To adjust for the potential bias, we applied adjustment parameters from a set of meta-analysis using the raw data extracted from the recent Cochrane review regarding test accuracy of MRI and TBx with or without SBx conditioning on PSA≥3 ng/mL ^9^ for strategy II. For strategy III and IV, we adjusted the test characteristics by taking the ISUP grading from STHLM-MR Phase I study ^15^ where patients undertook SBx on negative MRI (see Table 1 for input parameters of test characteristics; see Supplementary Material I Appendix A for strategy matrix of grading and details for adjustment).

**Table 1:**
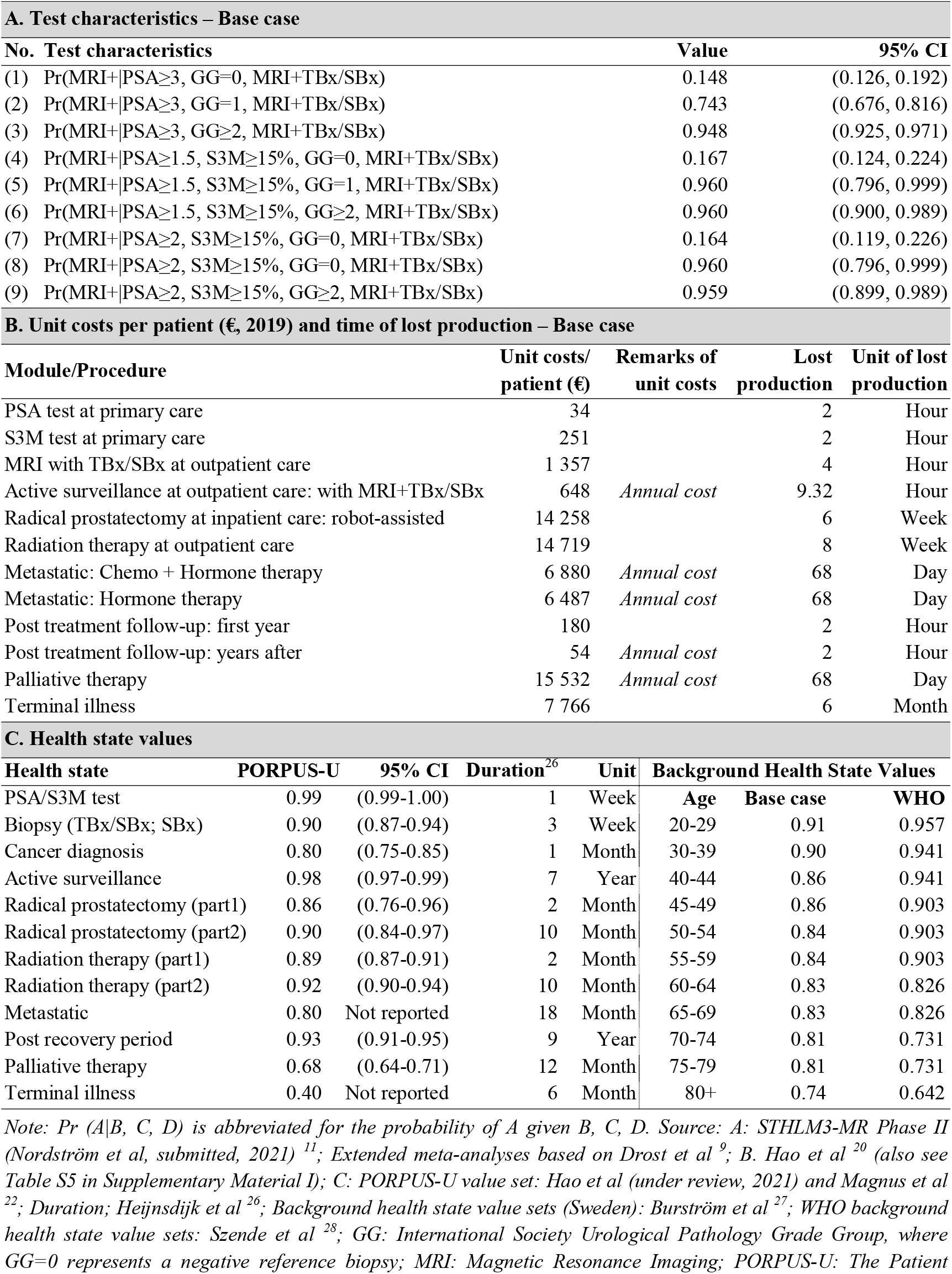
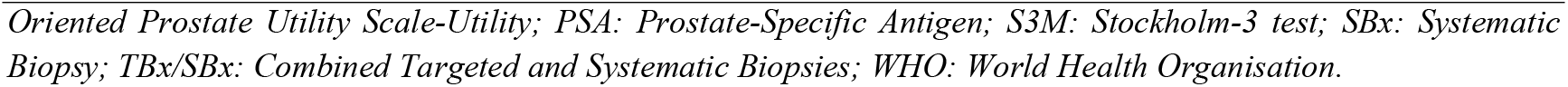
Input parameters used in the cost-effectiveness analysis

| A. Test characteristics – Base case |  |  |  |  |  |  |  |
| --- | --- | --- | --- | --- | --- | --- | --- |
| No. | Test characteristics | Value | 95% CI |  |  |  |  |
| (1) | Pr(MRI+\|PSA≥3, GG=0, MRI+TBx/SBx) | 0.148 | (0.126, 0.192) |  |  |  |  |
| (2) | Pr(MRI+\|PSA≥3, GG=1, MRI+TBx/SBx) | 0.743 | (0.676, 0.816) |  |  |  |  |
| (3) | Pr(MRI+\|PSA≥3, GG≥2, MRI+TBx/SBx) | 0.948 | (0.925, 0.971) |  |  |  |  |
| (4) | Pr(MRI+\|PSA≥1.5, S3M≥15%, GG=0, MRI+TBx/SBx) | 0.167 | (0.124, 0.224) |  |  |  |  |
| (5) | Pr(MRI+\|PSA≥1.5, S3M≥15%, GG=1, MRI+TBx/SBx) | 0.960 | (0.796, 0.999) |  |  |  |  |
| (6) | Pr(MRI+\|PSA≥1.5, S3M≥15%, GG≥2, MRI+TBx/SBx) | 0.960 | (0.900, 0.989) |  |  |  |  |
| (7) | Pr(MRI+\|PSA≥2, S3M≥15%, GG=0, MRI+TBx/SBx) | 0.164 | (0.119, 0.226) |  |  |  |  |
| (8) | Pr(MRI+\|PSA≥2, S3M≥15%, GG=0, MRI+TBx/SBx) | 0.960 | (0.796, 0.999) |  |  |  |  |
| (9) | Pr(MRI+\|PSA≥2, S3M≥15%, GG≥2, MRI+TBx/SBx) | 0.959 | (0.899, 0.989) |  |  |  |  |
| B. Unit costs per patient (€, 2019) and time of lost production – Base case |  |  |  |  |  |  |  |
| Module/Procedure | Unit costs/<br>patient (€) | Remarks of<br>unit costs | Lost<br>production | Unit of lost<br>production |  |  |  |
| PSA test at primary care | 34 |  | 2 | Hour |  |  |  |
| S3M test at primary care | 251 |  | 2 | Hour |  |  |  |
| MRI with TBx/SBx at outpatient care | 1 357 |  | 4 | Hour |  |  |  |
| Active surveillance at outpatient care: with MRI+TBx/SBx | 648 | Annual cost | 9.32 | Hour |  |  |  |
| Radical prostatectomy at inpatient care: robot-assisted | 14 258 |  | 6 | Week |  |  |  |
| Radiation therapy at outpatient care | 14 719 |  | 8 | Week |  |  |  |
| Metastatic: Chemo + Hormone therapy | 6 880 | Annual cost | 68 | Day |  |  |  |
| Metastatic: Hormone therapy | 6 487 | Annual cost | 68 | Day |  |  |  |
| Post treatment follow-up: first year | 180 |  | 2 | Hour |  |  |  |
| Post treatment follow-up: years after | 54 | Annual cost | 2 | Hour |  |  |  |
| Palliative therapy | 15 532 | Annual cost | 68 | Day |  |  |  |
| Terminal illness | 7 766 |  | 6 | Month |  |  |  |
| C. Health state values |  |  |  |  |  |  |  |
| Health state | PORPUS-U | 95% CI | Duration<sup>26</sup> | Unit | Background Health State Values |  |  |
| PSA/S3M test | 0.99 | (0.99-1.00) | 1 | Week | Age | Base case | WHO |
| Biopsy (TBx/SBx; SBx) | 0.90 | (0.87-0.94) | 3 | Week | 20-29 | 0.91 | 0.957 |
| Cancer diagnosis | 0.80 | (0.75-0.85) | 1 | Month | 30-39 | 0.90 | 0.941 |
| Active surveillance | 0.98 | (0.97-0.99) | 7 | Year | 40-44 | 0.86 | 0.941 |
| Radical prostatectomy (part1) | 0.86 | (0.76-0.96) | 2 | Month | 45-49 | 0.86 | 0.903 |
| Radical prostatectomy (part2) | 0.90 | (0.84-0.97) | 10 | Month | 50-54 | 0.84 | 0.903 |
| Radiation therapy (part1) | 0.89 | (0.87-0.91) | 2 | Month | 55-59 | 0.84 | 0.903 |
| Radiation therapy (part2) | 0.92 | (0.90-0.94) | 10 | Month | 60-64 | 0.83 | 0.826 |
| Metastatic | 0.80 | Not reported | 18 | Month | 65-69 | 0.83 | 0.826 |
| Post recovery period | 0.93 | (0.91-0.95) | 9 | Year | 70-74 | 0.81 | 0.731 |
| Palliative therapy | 0.68 | (0.64-0.71) | 12 | Month | 75-79 | 0.81 | 0.731 |
| Terminal illness | 0.40 | Not reported | 6 | Month | 80+ | 0.74 | 0.642 |

Note: Pr (A/B, C, D) is abbreviated for the probability of A given B, C, D. Source: A: STHLM3-MR Phase II (Nordström et al, submitted, 2021) <sup>11</sup>; Extended meta-analyses based on Drost et al <sup>9</sup>; B. Hao et al <sup>20</sup> (also see Table S5 in Supplementary Material I); C: PORPUS-U value set: Hao et al (under review, 2021) and Magnus et al <sup>22</sup>; Duration; Heijnsdijk et al <sup>26</sup>; Background health state value sets (Sweden): Burström et al <sup>27</sup>; WHO background health state value sets: Szende et al <sup>28</sup>; GG: International Society Urological Pathology Grade Group, where GG=0 represents a negative reference biopsy; MRI: Magnetic Resonance Imaging; PORPUS-U: The Patient
*Oriented Prostate Utility Scale-Utility; PSA: Prostate-Specific Antigen; S3M: Stockholm-3 test; SBx: Systematic Biopsy; TBx/SBx: Combined Targeted and Systematic Biopsies; WHO: World Health Organisation.*

Following a similar analytical approach to Karlsson et al ^6^, we simulated from the STHLM3-MR study and found the test characteristic thresholds on the PSA scale that corresponded to the relative test characteristics of the Stockholm3 test. For men with no cancer, ISUP GG=1 and GG≥2 cancers, we simulated for men who had a positive MRI at study entry. For those men, we chose the test characteristics threshold τ so that the number of men with PSA at or above τ was equal to the number of men who had a PSA≥3 ng/mL times the relative positive fraction for that group. Men with a PSA below the test characteristics threshold were assumed to be Stockholm3 negative. This approach assumes that the Stockholm3 test has similar prognostic characteristics as the PSA test.

### Reported outcomes

For each strategy, we modelled the mean lifetime number of screening tests, MRIs, biopsies, PCa incidence, mortality and life expectancy for 10^7^ males followed from age 55 years. We also reported PCa incidence during the screening ages 55-69 years.

### Cost-utility analysis

Based on the Swedish guidelines, the base-case cost-utility analysis was conducted from both a societal and healthcare perspective ^16^. The lifetime costs and QALYs were discounted at 3% per year ^16^ from age 55 years. ICERs were calculated as the difference in costs divided by the difference in QALYs between two interventions ^17^. We applied the categorical cost-effectiveness thresholds used by the National Board of Health and Welfare, where an ICER below 100,000 Swedish Krona (SEK) (1 Euro(€)=10.5892 SEK, 2019 ^18^, €9,444), between 100,000-499,999SEK (€9,444-47,218), between 500,000-1 Million (M) SEK (€47,218-€94,436) and above 1M SEK (€94,436) were defined as low, moderate, high and very high cost per QALY gained, respectively ^19^.

### Resource use and costs

Costs due to healthcare resources for screening, diagnosis, active surveillance, treatments, post-treatment follow-up and palliative care were considered as direct costs. Indirect costs of productivity losses due to morbidity were calculated using a human capital approach. Resource use and unit costs were taken from our previous study ^20^ and converted to the calendar year 2019 using the consumer price index ^21^ (see Table 1). For details of resource use and cost inputs, see Supplementary Material I Table S5.

### Health Outcomes

Health outcomes of each strategy were measured by QALYs, which were calculated by aggregating the product of health state values of individuals multiplied by the duration in each health state. The health state values were measured by the disease-specific Patient Oriented Prostate Utility Scale-Utility (PORPUS-U) (Hao et al, under review, 2021) based on a meta-analysis by Magnus et al ^22^. These values were multiplied by age-specific background health state values from the general population in Sweden measured by the generic instrument EQ-5D visual analogue scale (VAS) value set ^23^. We followed most health state durations from Heijnsdijk et al ^4^. Exceptions were metastatic disease for 18 months and palliative care for 12 months, based on the palliative register in Sweden (see Table 1).

### Sensitivity analyses

One-way sensitivity analyses were performed by varying: i) the threshold for a referral to MRI using S3M≥11% for strategy IV (note that we were unable to model for strategy III because the natural history model simulated for too few GG≥2 cancers) ; ii) the reflex threshold for Stockholm3 using PSA≥2.5 ng/mL for S3M≥11% and 15%, respectively; iii) the biopsy choice from the combined TBx/SBx to TBx alone on positive MRI for strategies III and IV (see test characteristics in Table 1 and Supplementary Material II Table S4); iv) the test characteristics calculated based on data from the STHLM3-MR using an ITT analysis, which included all patients in the experimental arm with ISUP grading from either TBx or SBx (for test characteristics used in (i) to (iv), see Supplementary Material II Table S4); v) the unit cost of Stockholm3 test, with a lower bound of €94 (1000 SEK) and a higher bound of €283 (3000 SEK); vi) the background health state value set measured for the Eur A countries from the World Health Organisation (WHO) measured by EQ-5D combined with an additional scale on recognition ^24^ (see Table 1); and vii) the discount rates to 0% and 5%.

A probabilistic sensitivity analysis included uncertainties in the relative positive fractions for Stockholm3, MRI test probabilities, costs and health state values. The Stockholm3 test characteristics were assumed to be normally distributed on a log scale. The MRI test probabilities as well as the health state values were assumed to be normally distributed on a logit scale. The costs were sampled from a gamma distribution of 95% confidence interval with ±20%. The model was assessed with the combination of new parameters 1000 times. We used cost-effectiveness acceptability curves (CEAC) to compare the screening strategies, presenting the possibility that a strategy being cost-effective relative to other alternatives at a given cost-effectiveness threshold. Furthermore, the expected value of perfect information (EVPI) was plotted to show the value that a policy maker would be willing to pay based on perfect information on all factors that influence the preference of choice by removing all uncertainties ^25^.

### Ethics

The use of the data from the Stockholm PSA and Biopsy Register was approved by the Ethical Review Board, Stockholm (dnr 2012/438-31/3, dnr 2016/620-32). The STHLM3-MR Study was approved by the Ethical Review Board, Stockholm (dnr 2017/1280-31) and the registration number was NCT03377881. All study participants have given written informed consent to publish these case details. All data were analysed anonymously.

## Results

### Base cases analysis

Compared with no screening, all screening strategies resulted in reductions of PCa deaths in a range of 7.0% to 8.6% and very similar QALY gains per man. Compared with PSA+MRI+TBx/SBx, using Stockholm3 as a reflex test and MRI+TBx/SBx for quadrennial screening reduced MRI by 50-60% and biopsies by 7-9% across a lifetime. From a societal perspective with a 3% discount rate, all screening strategies resulted in ICERs that were categorised as moderate costs per QALY gained in Sweden. PSA2+S3M15+MRI+TBx/SBx had the lowest ICER of €38,894 per QALY gained (see Table 2 and Figure 2).

**Table 2.** Summarised predictions in outcomes, costs and ICERs – Base case

| **Part A. Lifetime predictions in outcomes and costs for all strategies** |  |  |  |  |
| --- | --- | --- | --- | --- |
| **Lifetime predictions** | **Strategy I**<br>**No screening** | **Strategy II PSA3**<br>**+MRI+TBx/SBx** | **Strategy III PSA1.5+S3M15%**<br>**+MRI+TBx/SBx** | **Strategy IV PSA2+S3M15%**<br>**+MRI+TBx/SBx** |
| **Outcomes per 100,000 men** |  |  |  |  |
| MRI | 0 | 33 789 | 16 226 | 13 439 |
| Biopsy | 31 046 | 34 070 | 31 632 | 31 162 |
| Diagnosed PCa | 16 542 | 17 226 | 17 011 | 16 957 |
| Diagnosed PCa – GG≥2 | 11 907 | 12 201 | 12 186 | 12 156 |
| Diagnosed PCa (age 55-69) | 2 708 | 5 126 | 4 720 | 4 582 |
| Diagnosed PCa (age 55-69) – GG≥2 | 1 627 | 2 862 | 2 835 | 2 764 |
| Prostate cancer deaths | 5 774 | 5 280 | 5 342 | 5 370 |
| LY, undiscounted | 2 733 270 | 2 738 022 | 2 737 605 | 2 737 393 |
| QALYs, undiscounted | 2 203 907 | 2 206 800 | 2 206 640 | 2 206 523 |
| QALYS, discounted at 3% | 1 476 305 | 1 477 287 | 1 477 288 | 1 477 260 |
| QALYS, discounted at 5% | 1 182 630 | 1 183 048 | 1 183 087 | 1 183 083 |
| **Costs (M€) per 100,000 man** |  |  |  |  |
| Health care perspective, undiscounted | 362.6 | 395.5 | 401.7 | 395.2 |
| Health care perspective, discounted at 3% | 177.9 | 211.5 | 215.7 | 209.9 |
| Health care perspective, discounted at 5% | 116.1 | 148.1 | 151.5 | 146.3 |
| Societal perspective, undiscounted | 371.3 | 411.7 | 416.9 | 410.0 |
| Societal perspective, discounted at 3% | 184.3 | 224.4 | 227.6 | 221.5 |
| Societal perspective, discounted at 5% | 121.5 | 159.2 | 161.8 | 156.3 |
| **Part B. Incremental cost-effectiveness ratios (ICERs) – Societal perspective, 3% discounted for QALYs and costs** |  |  |  |  |
| **Strategy** | **Costs (M€) per**<br>**100,000 men** | **QALYs per**<br>**100,000 men** | **ICERs compared to** |  |
|  |  |  | **Lowest cost (No screening)** | **Next lowest cost** |
| No screening | 184.3 | 1 476 305 | - | - |
| PSA2+S3M15%+MRI+TBx/SBx | 221.5 | 1 477 260 | 38 894 | 38 894 |
| PSA3+MRI+TBx/SBx | 224.4 | 1 477 287 | 40 764 | 109 335 |
| PSA1.5+S3M15%+MRI+TBx/SBx | 227.6 | 1 477 288 | 43 993 | 2 061 150 |

*Note: the results are presented from age 55 years (conditional on survival to age 55). GG: International Society of Urological Pathology Grade Group; ICER: Incremental Cost-Effectiveness Ratio; LY: Life years; MRI: Magnetic Resonance Imaging; PCa: Prostate Cancer; PSA: Prostate-Specific Antigen; QALY: Quality-Adjusted Life-Year; SBx: Systematic Biopsy; S3M: Stockholm3 test; TBx/SBx: combined Targeted and Systematic Biopsies.*

**Figure 2:**
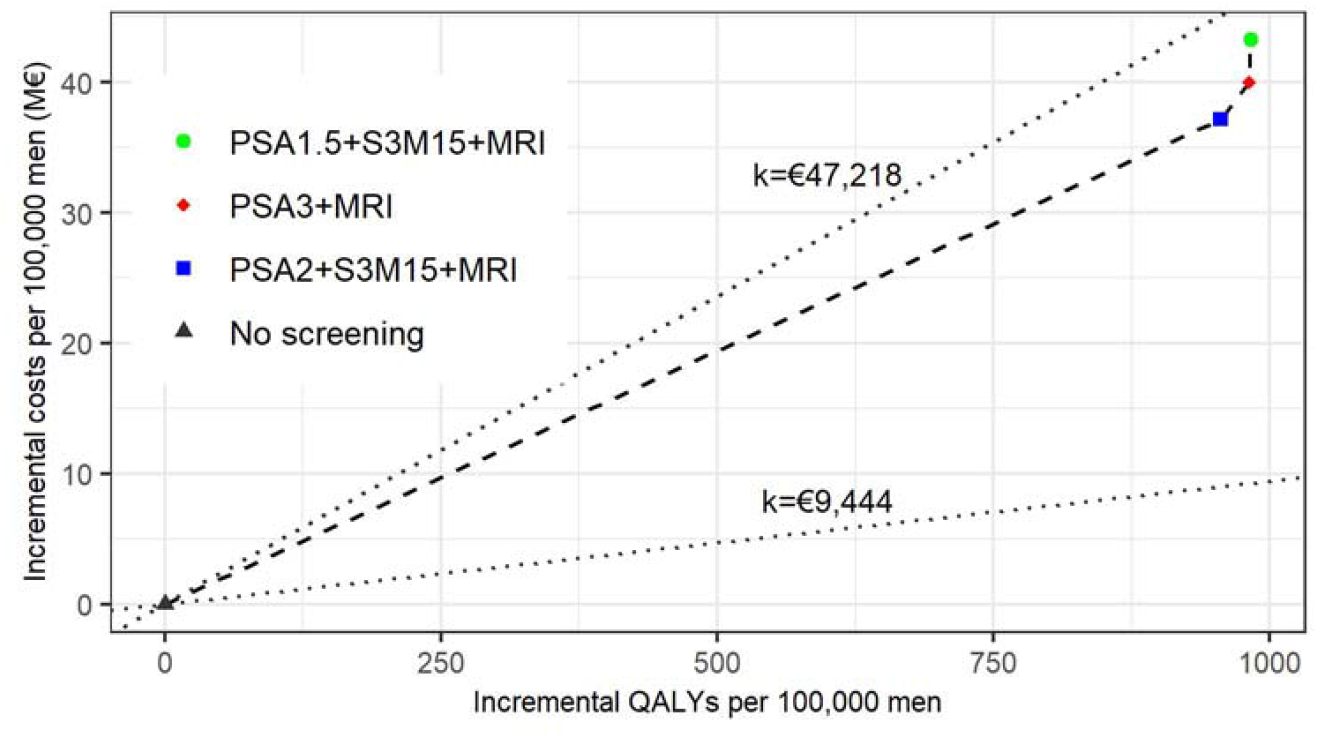
Cost-effectiveness plane: base case, societal perspective, 3% discounted. Legend: All screening strategies resulted in ICERs that were considered to be moderate cost per QALY gained in Sweden. The lower dashed line: cost-effectiveness threshold at €9,444/QALY (100,000SEK), which is considered to be low cost per QALY gained in Sweden; The upper dashed line: cost-effectiveness threshold at €47,218/QALY (500,000SEK), which is considered to be moderate cost per QALY gained in Sweden; The dashed line in the middle represents the cost-efficiency frontier. ICER: Incremental Cost-effectiveness Ratio; MRI: Magnetic Resonance Imaging; PSA: Prostate-Specific Antigen; QALY: Quality-Adjusted Life-Years; SEK: Swedish Krona; S3M: Stockholm3 test

Compared with PSA2+S3M15+MRI+TBx/SBx, PSA3+MRI+TBx/SBx had a very small gain in QALYs with an ICER over €109,335 per QALY gained, which is a very high cost in Sweden. Compared with PSA3+MRI+TBx/SBx, PSA1.5+S3M15+MRI+TBx/SBx had very similar QALYs gains and a marked increase in costs, resulting in an ICER over two million Euro.

### Sensitivity analysis

The results were robust in most of the one-way sensitivity analyses. The ICERs for the comparisons between the screening strategies and no screening continued to be categorised as moderate costs per QALY gained in Sweden. Reducing the unit cost of Stockholm3 to €94 (1000SEK), the PSA2+S3M15+MRI+TBx/SBx strategy showed an ICER that was 16% lower than in the base case. Applying background health state values reported by WHO increased the ICERs by over 20% for all screening strategies. The results were sensitive to the discount rates (see Figure 3).

**Figure 3:**
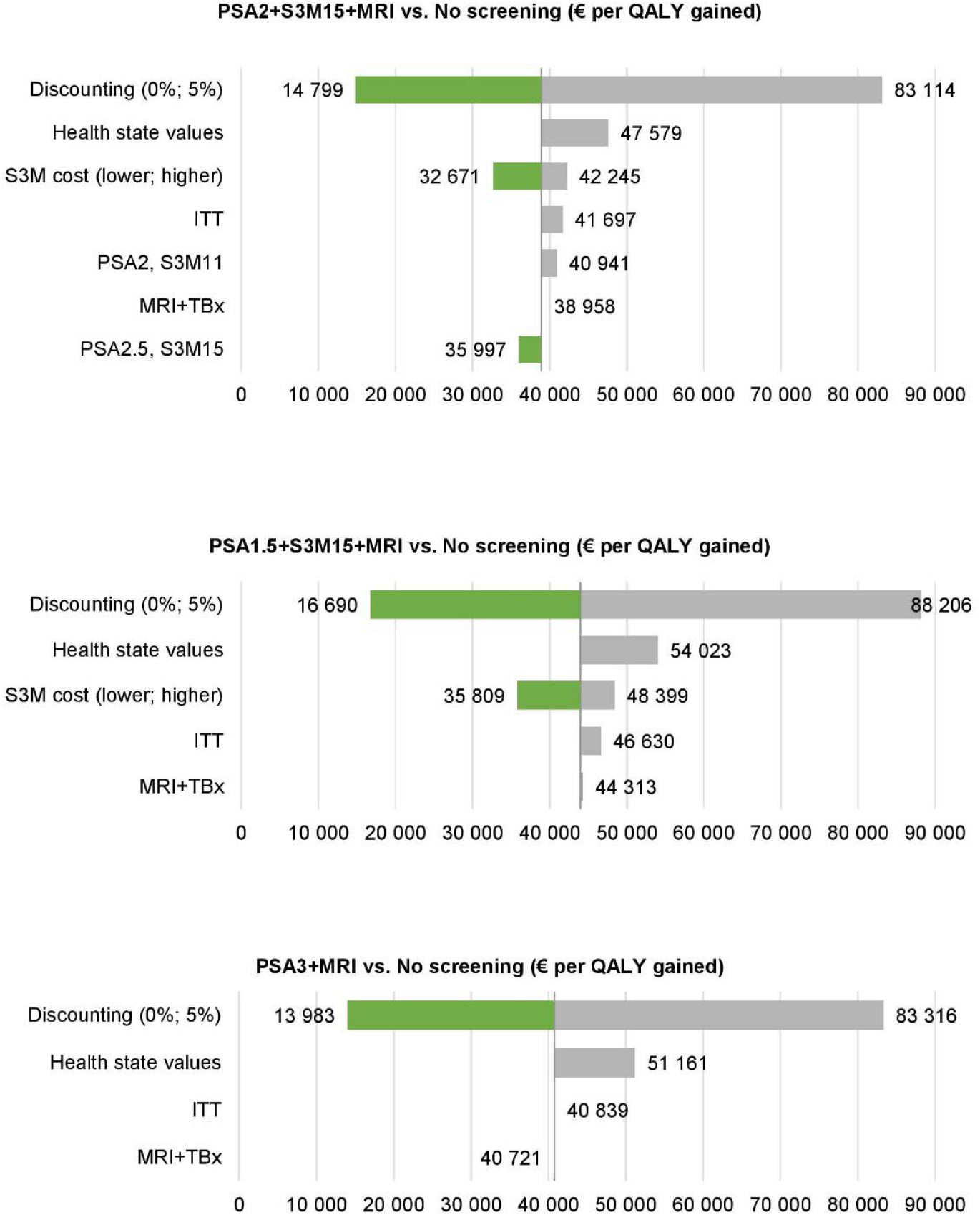
One-way sensitivity analysis in tornado plot.

ICERs from the one-way sensitivity comparing screening strategies with no screening, using: 0% and 5% discount rates; background health state values reported by the World Health Organisation; test characteristics using reflex threshold of PSA≥2ng/mL for S3M>=11%; test characteristics by the ITT method; MRI with TBx instead of MRI and TBx/SBx; and lower S3M cost at 94€ (1000SEK) and higher S3M cost at €283 (3000SEK).

ICER: Incremental Cost-effectiveness Ratio; MRI: Magnetic Resonance Imaging; PSA: Prostate-Specific Antigen; QALY: Quality-Adjusted Life-Years; SEK: Swedish Krona; S3M: Stockholm3 test; TBx: Targeted Biopsy; TBx/SBx: combined Targeted and Systematic Biopsies

The cost-effectiveness acceptability curve (see Figure 4) showed that using a cost-effectiveness threshold of €47,218 (SEK 500,000) per QALY gained, the probability that no screening would be cost-effective was 0%, whereas PSA2+S3M15+MRI was expected to be cost-effective at a probability of 70%. At the same threshold, the expected value of perfect information was close to €0.5M per 100,000 men (see Supplementary Material I Figure S3). The EVPI exhibited the first peak at a cost-effectiveness threshold of €40,000 per QALY gained, where the optimal strategy changed from no screening to PSA2+S3M15+MRI on the CEAC. At a threshold of €83,000 per QALY gained, the probabilities that PSA+MRI and PSA2+S3M15+MRI would be cost-effective were equal.

**Figure 4:**
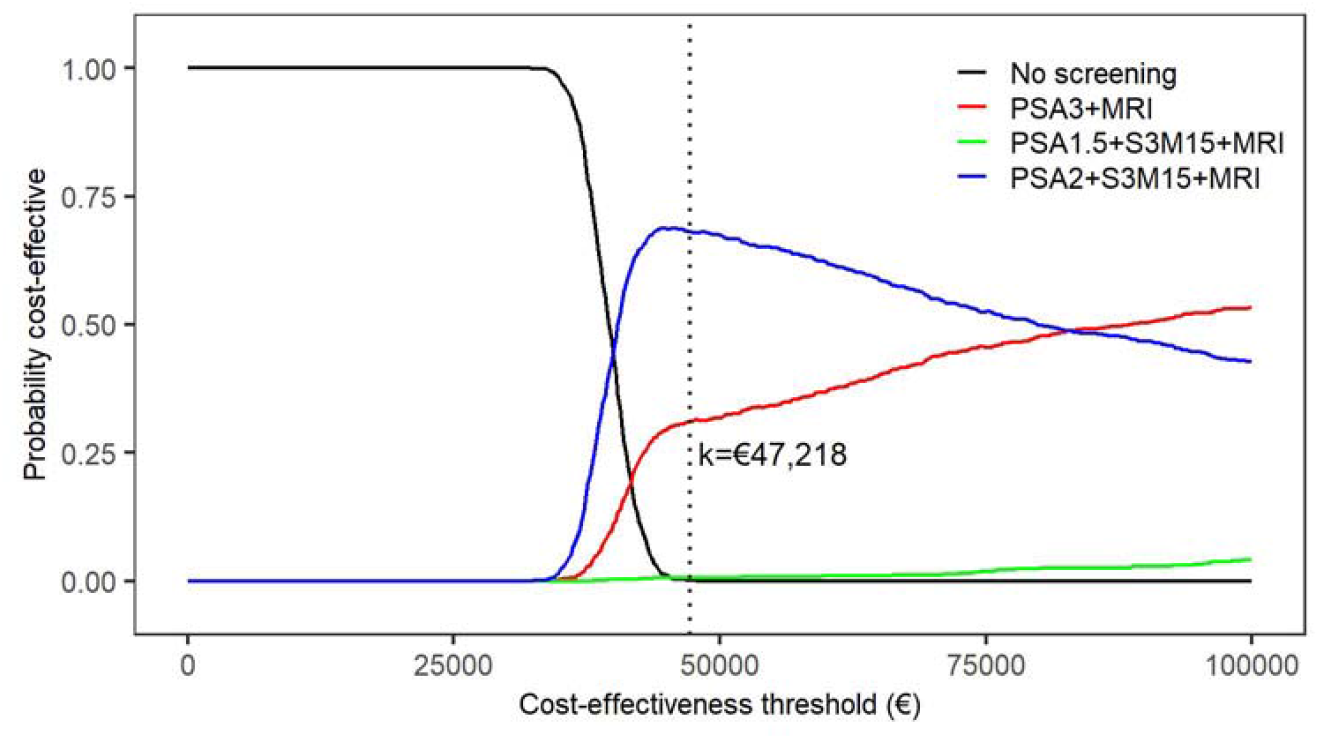
Cost-effectiveness acceptabililty curve. MRI: Magnetic Resonance Imaging; PSA: Prostate-Specific Antigen; QALY: Quality-Adjusted Life-Years; S3M: Stockholm3 test.

## Discussion

### Main findings

In summary, the model predicted that the three MRI-based screening strategies reduced PCa related deaths by 7% to 8.6% and resulted in ICERs below €47,218 per QALY gained compared with no screening, which is considered to be a moderate cost per QALY gained in Sweden. These results were sensitive to the background health state values reported by WHO and discount rates. Stockholm3 with a reflex threshold of PSA≥2ng/mL had the lowest ICER, €38,894 per QALY gained, in the base case analysis. Reducing the unit cost of Stockholm3 to €94 (57% reduction) resulted in a 16% reduction in the ICER. In relation to screening with PSA, Stockholm3 with a reflex threshold of PSA≥2ng/mL could reduce the use of MRI by 60% and further reduce the number of biopsies by 9% across a lifetime. Compared with MRI-based screening with Stockholm3 at a reflex threshold of PSA≥2ng/mL, screening with PSA resulted in an ICER that was over €100,000 per QALY gained, which was considered to be a very high cost in Sweden. At a cost-effectiveness threshold of €47,218 per QALY gained, the probability that the MRI-based screening with Stockholm3 at a reflex threshold of PSA≥2ng/mL would be cost-effective compared to the other strategies was 70%.

### Comparison with existing studies and interpretation of selected results

To our knowledge, no existing study has assessed the cost-effectiveness of prostate cancer screening using PSA and another triage diagnostic test in combination with MRI. One study found that adding the Prostate Health Index (PHI) to MRI was associated with improved predictions of overall and high risk cancers of GG≥2 ^29^. Another study also concluded that PHI could be effective in reducing MRIs and biopsies without compromising the detection of GG≥2 cancers ^30^. However, these studies were not conducted in a screening context. Other blood tests may be competitive alternatives to Stockholm3, however lack of evidence makes the comparison difficult.

The ICERs of the screening strategies compared with no screening in this study lay in the categories of moderate cost per QALY gained in Sweden. Our assessment of the cost-effectiveness of PSA and MRI compared with no screening is qualitatively different from our prediction based on the Cochrane review (Hao et al, under review 2021), where we now estimate an ICER that is approximately 50% lower (€40,764 vs €85,001). This change may be explained by a difference in the probability of positive MRI given true negative of benign biopsies (GG=0). For biopsy-naïve men, the proportion of a negative MRI result from the STHLM3-MR screening trial given PSA≥3ng/mL was 62% (Nordström et al, submitted, 2021) ^11^, twice of the proportion estimated from the Cochrane review (31%) ^9^. As MRI remains a resource-intensive diagnostic approach and is highly dependent on the operators ^30^, this further implied that radiologists are less likely to over-grade the PI-RADS in a screening setting. We also noticed that the outcomes per 100,000 men in this study for the number of biopsies, PCa diagnosis and deaths were slightly higher than our previous predictions (Hao et al, under review 2021), which are due to the updated deaths rate from the Human Mortality Database being lower ^31^.

In addition, the cost of biopsy in Sweden may be comparatively high compared with other European countries. In the Netherlands, the diagnosis including a systematic biopsy, a pathological research and a GP consultation costs approximately €240 ^32^ whilst a diagnosis including an urologist and nurse visit, a systematic biopsy and a pathological assessment costs was almost four-fold higher (€875) in Sweden. If the biopsy costs in Sweden were reduced, then the ICERs comparing the screening strategies with no screening would be further reduced.

Our base case analysis was per protocol, whereas Nordström et al ^11^ used the ITT method. Using the test characteristics by ITT from the STHLM3-MR study showed very similar results to our base case analysis. It is difficult to model the ITT non-compliance because the disease status is unknown for those who did not comply, whereas a per-protocol analysis would assess the efficacy for those who did comply.

### Strengths and limitations

This is the first study to investigate the cost-effectiveness of the combination of PSA, Stockholm3 and MRI compared with PSA and MRI for risk-stratified screening of prostate cancer based on the evidence from the STHLM3-MR study (Nordström et al, submitted, 2021) ^11^ and the recent Cochrane review ^9^. To our knowledge, none of the existing studies have assessed the cost-effectiveness of a reflex test in addition to PSA and MRI within a screening context. Another strength of this study is that the natural history model has been carefully calibrated with good internal validity for comparisons of screening interventions. It allows for stage shift to provide a more accurate lifetime representation of the effects of the screening, diagnostic and treatment processes. In addition, the health state values were available from our recent study (Hao et al, under review, 2021) using consistent outcome measures. Moreover, the costs along the screening and disease management pathways in Sweden were based on the results from our recent study of the societal cost of PCa ^20^.

Some limitations should be noted. First, per the STHLM3-MR study protocol, patients with a negative MRI result but S3M≥25% were referred to undertake a SBx. We did not include these strategies into our analysis due to the comparatively poor test characteristics. Importantly, such a high-risk strategy was intended to complement PI-RADS and reduce potential up-staging due to safety concerns. An alternative high-risk strategy, such as referring all men with PSA≥10 ng/mL to biopsy, would support the introduction of MRI in the diagnostic pathway. Second, the estimated STHLM3-MR test characteristics had comparatively wide confidence intervals. Further studies would increase the precision for pooled contrasts between PSA and PSA2+S3M. Third, we did not evaluate how different screening intervals (e.g. 2-, 3- or 5-yearly) would affect our findings. However, our earlier study predicted that 4-yearly screening was considered as an optimal choice among screening strategies (Hao et al, under review, 2021). Finally, as PCa incidence, mortality rates and costs in Sweden may differ from or be comparatively higher than other countries, our results may not be generalisable to other populations.

### Policy implications

Compared with no screening, using Stockholm3 test at a reflex threshold of 2ng/mL and a test threshold of S3M≥15% with MRI and combined TBx/SBx resulted in a moderate cost per QALY gained in Sweden and may be considered cost-effective. This strategy was associated with a reduction in PCa deaths and could be considered as a risk-stratification screening strategy. Any reduction in the unit cost of Stockholm3 would further improve the cost-effectiveness of this test for early detection of PCa. In the future, it would also be useful to consider a different age range (e.g. 50-74).

The evidence from this study can support the development and evaluation of several pilot projects of organised prostate cancer testing currently underway in Sweden. Swedish policy-makers can then make a decision whether a national programme for organised prostate cancer testing could reduce the harms from PSA testing.

## Conclusion

Compared with no screening, use of PSA test or Stockholm3 as a reflex test with MRI and combined TBx/SBx were predicted to result in ICERs below €47,218 per QALY gained, which is considered a moderate cost per QALY gained in Sweden. The use of Stockholm3 test at a reflex threshold of PSA≥2ng/mL was predicted to have very similar QALY gains but lower costs to other screening strategies and a 60% reduction in the number of MRI compared to PSA. Thus, Stockholm3 at a reflex threshold of PSA≥2ng/mL with MRI and combined TBx/SBx can be considered a cost-effective screening strategy for prostate cancer that reduces screening-related harms and costs while maintaining health benefits from early detection.

## Supporting information

Supplementary Material III

## Data Availability

The raw data used for simulation from the Stockholm PSA and Biopsy Register (SPBR) are potentially identifiable and data access to those data has been restricted by the Stockholm Ethical Review Board. The authors do not own these data and had no special privileges in accessing the data. Anyone wishing to access the individual level data would need to apply for permission through an Ethical Review Board and from the primary data owners, including the Swedish National Board of Health and Welfare and Karolinska Institutet.

https://github.com/mclements/prostata

## Acknowledgements

We are grateful for data management of the Stockholm PSA and Biopsy Register by Pouran Almstedt.

## References

1. World Health Organisation. Global Cancer Observatory, Cancer Overtime, WHO Cancer Mortality Database. 20 June 2019. https://www-dep.iarc.fr/WHOdb/WHOdb.htm. Accessed 09 Nov 2020.

2. Hugosson J, Roobol MJ, Mansson M, et al. A 16-yr follow-up of the European Randomized study of Screening for Prostate Cancer. Eur Uro. 2019;76(1):43–51.

3. Heijnsdijk EA, der Kinderen A, Wever EM, et al. Overdetection, overtreatment and costs in prostate-specific antigen screening for prostate cancer. Br J Cancer. 2009;101(11):1833–1838.

4. Heijnsdijk EA, de Carvalho TM, Auvinen A, et al. Cost-effectiveness of prostate cancer screening: a simulation study based on ERSPC data. J Natl Cancer Inst. 2015;107(1):366.

5. Gronberg H, Adolfsson J, Aly M, et al. Prostate cancer screening in men aged 50-69 years (STHLM3): a prospective population-based diagnostic study. Lancet Oncol. 2015;16(16):1667–76.

6. Karlsson AA, Hao S, Jauhiainen A, et al. The cost-effectiveness of prostate cancer screening using the Stockholm3 test. PloS One. 2021;16(2):e0246674.

7. Regionala Cancercentrum i Samverkan. Prostatacancer Nationellt vårdprogram. 2020. https://www.cancercentrum.se/globalassets/cancerdiagnoser/prostatacancer/vardprogram/nationellt-vardprogram-prostatacancer.pdf. Accessed 2020-11-09.

8. European Association of Urology. Oncology guidelines prostate cancer. Edn. presented at the EAU Annual Congress Amsterdam 2020. ISBN 978-94-92671-07-3. 2020. EAU Guidelines. https://uroweb.org/guideline/prostate-cancer/#1

9. Drost FH, Osses DF, Nieboer D, et al. Prostate MRI, with or without MRI-targeted biopsy, and systematic biopsy for detecting prostate cancer. Cochrane Database Syst Rev. 2019;4:Cd012663.

10. Schoots IG, Roobol MJ, Nieboer D, et al. Magnetic resonance imaging-targeted biopsy may enhance the diagnostic accuracy of significant prostate cancer detection compared to standard transrectal ultrasound-guided biopsy: a systematic review and meta-analysis. Eur Urol. 2015;68(3):438–450.

11. Nordström T, Discacciati A, Bergman M, et al. Prostate cancer screening using a combination of risk-prediction, magnetic resonance imaging and targeted prostate biopsies: a randomised trial. doi:http://dx.doi.org/10.2139/ssrn.3788918 https://ssrn.com/abstract=3788918

12. Regionala Cancercentrum i Samverkan. Rekommendationer om organiserad prostatacancertestning (OPT) (2020).

13. Karlsson A, Jauhiainen A, Gulati R, et al. A natural history model for planning prostate cancer testing: Calibration and validation using Swedish registry data. PLoS One. 2019;14(2):e0211918.

14. Nordström T, Aly M, Clements M, Weibull C, Adolfsson J, Grönberg H. Prostate-specific Antigen (PSA) testing is pevalent and increasing in Stockholm County, Sweden, despite no recommendations for PSA screening: results from a population-based study, 2003-2011. Eur Uur. 2013;63(3):419–425.

15. Nordstrom T, Picker W, Aly M, et al. Detection of prostate cancer using a multistep approach with prostate-specific antigen, the Stockholm 3 test, and targeted biopsies: The STHLM3 MRI Project. Eur Urol Focus. 2017;3(6):526–528.

16. Tandvårds-och Läkemedelsförmånsverket. Tandvårds-och Läkemedelsförmånsverkets Allmänna Råd. TLVAR 2017:1 Ändring i Tandvårds-och Läkemedelsförmånsverkets Allmänna råd (TLVAR 2003:2) om Ekonomiska Utvärderingar. In. Stockholm 2017.

17. Drummond MF, Sculpher MJ, Claxton K, et al. Methods for the Economic Evaluation of Health Care Programmes (4^th^ ed). New York: Oxford University Press, 2015.

18. Riksbanken. Annual average exchange rates. https://www.riksbank.se/en-gb/statistics/search-interest--exchange-rates/annual-average-exchange-rates/. Accessed 2020-12-02.

19. Nationella Riktlinjer för Hjärtsjukvård. Hälsoekonomiskt Underlag Bilaga. (2018).

20. Hao S, Östensson E, Eklund M, et al. The economic burden of prostate cancer – a Swedish prevalence-based register study. BMC Health Serv Res. 2020;20(1):448.

21. Statistikmyndigheten. CPI, Fixed Index Numbers (1980=100). Statistikmyndigheten. https://www.scb.se/en/finding-statistics/statistics-by-subject-area/prices-and-consumption/consumer-price-index/consumer-price-index-cpi/pong/tables-and-graphs/consumer-price-index-cpi/cpi-fixed-index-numbers-1980100/. Updated 2020-11-12. Accessed 2020-12-02

22. Magnus A, Isaranuwatchai W, Mihalopoulos C, et al. A systematic review and meta-analysis of prostate cancer utility values of patients and partners between 2007 and 2016. MDM Policy Pract. 2019;4(1): 2381468319852332.

23. Burström K, Johannesson M, Diderichsen F. Swedish population health-related quality of life results using the EQ-5D. Qual. Life Res. 2001;10(7):621–35.

24. World Health Organisation. Cost effectiveness and strategic planning (WHO-CHOICE). Health State Valuations. https://www.who.int/choice/demography/health_valuations/en/. Published 2020. Accessed May 16 2020.

25. Expected Value of Perfect Information (EVPI). York Health Economics Consortium. https://yhec.co.uk/glossary/expected-value-of-perfect-information-evpi/. Accessed January 28 2021

26. Heijnsdijk EA, Wever EM, Auvinen A, et al. Quality-of-life effects of prostate-specific antigen screening. The N Engl J Med. 2012;367(7):595–605.

27. Burström K, Rehnberg C, Hälsoekonomi CffEfso. Hälsorelaterad livskvalitet i Stockholms län 2002: resultat per åldersgrupp och kön, utbildningsnivå, födelseland samt sysselsättningsgrupp : en befolkningsundersökning med EQ-5D. Centrum för folkhälsa; 2006.

28. EQ-5D index population norms (European VAS value set). In: Szende A, Janssen B, Cabases J, eds. Self-reported population health: an international perspective based on EQ-5D. Springer. Copyright 2014, Szende, A., Janssen, B., Cabases, J.; 2014.

29. Gnanapragasam VJ, Burling K, George A, et al. The Prostate Health Index adds predictive value to multi-parametric MRI in detecting significant prostate cancers in a repeat biopsy population. Sci Rep. 2016;6:35364.

30. Kim L, Boxall N, George A, et al. Clinical utility and cost modelling of the phi test to triage referrals into image-based diagnostic services for suspected prostate cancer: the PRIM (Phi to RefIne Mri) study. BMC Med. 2020;18(1):95.

31. Human Mortality Database. University of California, Berkeley (USA), and Max Planck Institute for Demographic Research (Germany); 2020. https://www.mortality.org/. Accessed December 8 2020.

32. Getaneh AM, Heijnsdijk EAM, Roobol MJ, et al. Assessment of harms, benefits, and cost-effectiveness of prostate cancer screening: A micro-simulation study of 230 scenarios. Cancer Med. 2020;9(20):7742–7750.

