## Supplementary Material I for "Cost-effectiveness of Stockholm3 test and magnetic resonance imaging in prostate cancer screening: a microsimulation study"

**Supplementary Materials I**

**Figure S1 Natural history simulation model**

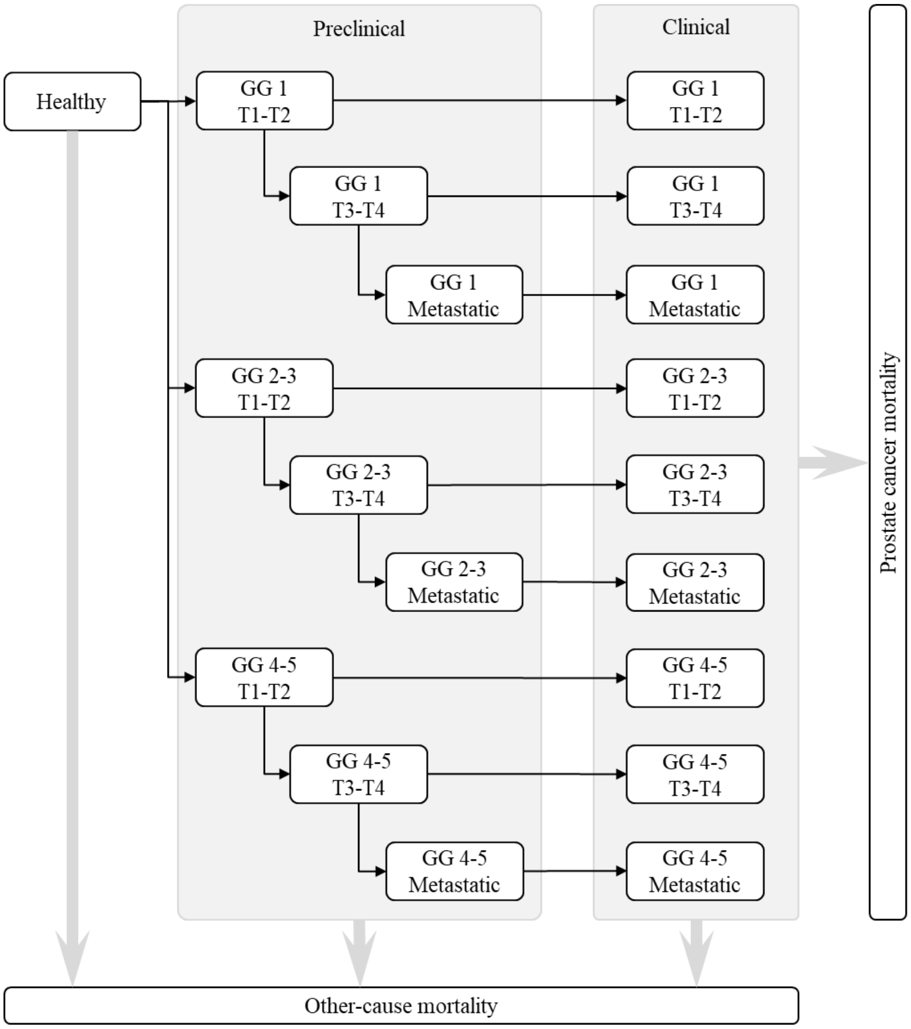

The model reflects cancer onset by ISUP (GG 1; GG 2-3; GG 4-5), with progression by T-stage and to metastatic cancer. Preclinical cancers may be clinically diagnosed, with cause-specific survival from the time of clinical diagnosis. Death due to other causes is represented as a competing event. ISUP: International Society of Urological Pathology: GG: grade group

### Appendix A. Test characteristics parameter adjustment

#### A1 Methods

For the adjustment parameters used for the strategy I using prostate-specific antigen (PSA) test, magnetic resonance imaging (MRI) and combined targeted and systematic biopsy (TBx/SBx), we reviewed the 25 papers used in the Agreement analysis from Drost (2019) [1] and extracted data for the extended meta-analysis. The studies used for the agreement analysis Strategies are conditional on biopsy naïve patients with PSA≥3 (Table S1).

For the adjustment parameters used for Strategy III and IV, which were using TBx/SBx on patients with positive MRI given prostate-specific antigen (PSA)≥1.5ng/mL and Stockholm3 (S3M) test≥15% and given with PSA≥2ng/mL and S3M≥15%, respectively, we used the data from STHLM3-MR Phase I study which patients with negative MRI results also undertook SBx [2]. This would provide additional information on the true negative and true positive fractions.

For the test parameter Pr(MRI+|PSA+, GG=0, MRI+TBx/SBx, we calculated the adjusted 95% confidence interval (CI) using following information:

*p1 = y1/n1, p2 = y2/n2, ratio = p1/p2*

*meanlog=log(ratio), varlog=(1-p1)/p1/n1+(1-p2)/p2/n2*

*lower=meanlog-1.96*sqrt(varlog), upper=meanlog+1.96*sqrt(varlog)*

where y1 represents the number of benign cancers on MRI+ detected by TBx/SBx and n2 represents the total number of negative MRI results (MRI-) and the number of benign cancers on MRI+ detected by TBx/SBx from the STHLM3-MR study; y2 represents the number of benign cancers on MRI+ detected by TB/SBx and benign cancers detected by SBx on MRI- using the Cochrane Review [1]; n2 represents the number of benign cancers on MRI+ detected by TBx/SBx and number of MRI- results from the Cochrane Review [1].

For the test parameters Pr(MRI+|PSA+, GG=1, MRI+TBx/SBx) and Pr(MRI+|PSA+, GG⩾2, MRI+TBx/SBx), the parameter values were equal to 1 by the strategy matrix described (see A1.2). We used the 95% CI results directly from the meta-analysis of the adjusted parameters.

For the test characteristics Pr(MRI+|PSA≥1.5, S3M≥15%, GG=0, MRI+TBx/SBx) and Pr(MRI+|PSA⩾2, S3M⩾ 15%, GG=0, MRI+TBx/SBx), we calculated the adjusted 95% CI using following information:

*p1 = y1/n1, p2 = y2/n2, ratio = p1/p2*

*meanlog=log(ratio), varlog=(1-p1)/p1/n1+(1-p2)/p2/n2*

*lower=meanlog-1.96*sqrt(varlog), upper=meanlog+1.96*sqrt(varlog)*

where y1 represents the number of benign cancers on positive PSA and S3M test and MRI+ detected by TBx/SBx and n2 represents the total number of negative MRI results (MRI-) and the number of benign cancers on positive PSA and S3M tests and MRI+ detected by TBx/SBx from the STHLM3-MR study; y2 represents the number of benign cancers on positive PSA and S3M test and MRI+ detected by TB/SBx and benign cancers detected by SBx on MRI- using the STHLM3-MR Phase I study [2]; and n2 represents the number of benign cancers on positive PSA and S3M tests and MRI+ detected by TBx/SBx and number of MRI- results from the STHLM3-MR Phase I study [2].

For the rest test characteristics in the base case including Pr(MRI+|PSA≥1.5, S3M≥15%, GG=1, MRI+TBx/SBx), Pr(MRI+|PSA≥1.5, S3M≥15%, GG≥2, MRI+TBx/SBx), Pr(MRI+|PSA≥2, S3M≥15%, GG=1, MRI+TBx/SBx) and Pr(MRI+|PSA≥2, S3M≥15%, GG≥2, MRI+TBx/SBx), the parameter values were equal to 1 given the strategy matrix described (see A1.2). We used 95% CI results from the binomial tests for the adjusted parameters, based on the proportion of GG=1 or GG≥2 cancers detected by TBx/SBx on MRI+ to the total number of GG=1 or GG≥2 cancers detected by SBx on MRI- and TBx/SBx on MRI+ from the STHLM3-MR Phase I study [2].

**A1.1 Selection criteria of the studies for meta-analyses**

Based on the summarised characteristics of the agreement studies in Drost et al. [1], we excluded Boesen (2017), Jambor (2017) and Costa (2013) which had insufficient data. We also excluded Petier (2015) which was unable to differentiate the number of patients with negative biopsies between MRI- and MRI+, Delongchamps (2013) which did not have details on the number of patients for GG=1 and GG≥2, and Chen (2015) which only focused on patients with GG≥2. In addition, Okcelik (2016) which used PCA3 prior to MRI with a 0-1 scale was excluded. Table S1 summarised the study characteristics.

For the adjustment parameters used for biopsy-naïve patients, in total, 16 studies were included for the meta-analyses. Twelve studies used MRI scoring with PI-RADS scale 1-5, of which 10 studies used PI-RADS≥3 as a threshold for MRI+ and two studies used PI-RADS≥4 as threshold for MRI+ (Jambor (2015) and Kim (2017)). Lee (2016), Lee

(2017) and Tonttilla (2016) took MRI scoring with a scale of 1-4 and score 2 as the threshold for MRI+. A different type of scale of MRI scoring was used by Cool (2016).

Table S1 Study characteristics

| **Study** | **Setting** | **MRI scale; Threshold** | **MRI TBx Technique** | **No. Cores** MRI TBx | **No. Cores** SBx | **Participants** Biopsy naive | **Median age** (Range/SD) | **Median PSA (ng/ml)** (Range/SD) |
|---|---|---|---|---|---|---|---|---|
| **Biopsy-naïve patients** | | | | | | | | |
| Albert (2017) | Netherlands | 1-5, ≥3 | Software | 2 cores/lesion | 6/12 (Group1/2,3) | 74 | 73 (72-74) | 4.2 (3.4-5.8) |
| Boesen (2018) | Denmark | 1-5, ≥3 | Software | 1-2 cores/lesion | 10-core | 1020 | 67 (61-71) | 8 (5.7-13) |
| Castellucci (2017) | Spain | 1-5, ≥3 | Cognitive | 2 cores | Not reported | 168 | 61 (8) | 8.3 (6.1) |
| Cool (2016) | Canada | Other | Software | 1-3 cores/lesion | 12-core+2 cores | 50 | 59 (8) | 6 (3.5) |
| Filson (2016) | USA | 1-5, ≥3 | Software | 1 core, longest region | 12-core | 329 | 64 (59-69) | 5.8 (4.4-8.1) |
| Garcia Bennett (2017) | Spain | 1-5, ≥3 | Cognitive | Average 1.8 cores | 12-core | 60 | 64 (6.7) | 7.2 (6-9.4) |
| Grönberg (2018) | Sweden, Norway | 1-5, ≥3 | Software | 2-3 cores/lesion | 12-core | 387 | 64 (45-74) | 6.3 (4.4) |
| Jambor (2015) | Finland, Slovakia | 1-5, ≥4 | Cognitive | 5 cores | 12-core | 53 | 66 (47-76) | 7.4 (4-14) |
| Kim (2017) | USA | 1-5, ≥4 | Software, Cognitive | Not reported | 12-core | 183 | 64 (7) | 10.2 (15.1) |
| Lee (2016) | Korea | 1-4, ≥2 | Cognitive | 1 core/lesion | 12-core | 76 | 66 (43-83) | 6.3 (3.3-9.8) |
| Lee (2017) | Korea | 1-4, ≥2 | Cognitive | Not reported | 12-core | 123 | 62 (10) | 6.4 (1.8) |
| Panebianco (2015) | Italy | 1-5, ≥3 | Cognitive | 12-core TBx+SBx | 14-core | 570 | 64 (51-82) | Not reported |
| Pokorny (2014) | Australia | 1-5, ≥3 | In-bore | 2-3 cores/lesion | 12-core | 223 | 63 (57-68) | 5.3 (4.1-6.6) |
| Rouvière (2019) | France | 1-5, ≥3 | Software | 1-2 cores/lesion | 12-core | 251 | 64 (59-68) | 6.5 (5.6-9.6) |
| Tonttilla (2016) | Finland | 1-4, ≥2 | Cognitive | 1-2 cores/lesion; max. 2 lesions | 10 to 12-core | 53 | 63 (60-66) | 6.1 (4.2-9.9) |
| Van der Leest (2018) | Netherlands | 1-5, ≥3 | In-bore | 2-4 cores | 12-core | 626 | 65 (59-68) | 6.4 (4.6-8.2) |

Source: Drost et al [1]. There are three common techniques to guide the TBx: i) cognitively, where the urologist has knowledge of the location of the targeted lesion from the MRI; ii) fusion, where the MRI is fused with the real-time ultrasound images to target the lesion; and iii) the direct in-bore technique, where a patient's biopsy is undertaken in a MRI suite, allowing for real-time MR-imaging and lesion targeting [3,4]. However, there is little evidence that any technique is superior over another [3]. MRI: Magnetic Resonance Imaging; PSA: Prostate-Specific Antigen; SBx: Systematic Biopsy; SD: Standard Deviation; TBx: Targeted Biopsy; USA: United States of America

### A1.2 Strategy matrix

#### Strategy matrix PSA+MRI+TBx/SBx

| | MRI- | MRI+TBx | | | |
|---|---|---|---|---|---|
| SBx | | GG=0 | GG=1 | GG≥2 | SBx Total |
| GG=0 | x | a | d | g | S0 |
| GG=1 | y | b | e | h | S1 |
| GG≥2 | z | c | f | i | S2 |
| **MRI Total** | **M0** | **T0** | **T1** | **T2** | |

GG* definition

GG*=0 GG*=1 GG*≥2

MRI+, TBx- | GG*=0

MRI+, TBx+, SBx+ | GG*=1

MRI+, TBx+, SBx+ | GG*≥2

#### Strategy matrix S3M+MRI+TBx/SBx

| | MRI- | MRI+TBx | | | |
|---|---|---|---|---|---|
| SBx | | GG=0 | GG=1 | GG≥2 | SBx Total |
| GG=0 | x' | a | d | g | S0 |
| GG=1 | y' | b | e | h | S1 |
| GG≥2 | z' | c | f | i | S2 |
| **MRI Total** | **M0** | **T0** | **T1** | **T2** | |

GG* definition

GG*=0 a+M0-y'-z' GG*=1 b+d+e+y' GG*≥2 c+f+g+h+i+z'

MRI+, TBx- | GG*=0

MRI+, TBx+, SBx+ | GG*=1

MRI+, TBx+, SBx+ | GG*≥2

#### Strategy matrix PSA+MRI+TBx

| | MRI- | MRI+TBx | | | |
|---|---|---|---|---|---|
| SBx | | GG=0 | GG=1 | GG≥2 | **SBx Total** |
| GG=0 | x | a | d | g | **S0** |
| GG=1 | y | b | e | h | **S1** |
| GG≥2 | z | c | f | i | **S2** |
| **MRI Total** | **M0** | **T0** | **T1** | **T2** | |

GG* definition

GG*=0 GG*=1 GG*≥2

MRI+, TBx- | GG*=0

MRI+, TBx+ | GG*=1

MRI+, TBx+ | GG*≥2

**Strategy matrix S3M+MRI+TBx**

| | MRI- | MRI+TBx | | | |
|---|---|---|---|---|---|
| SBx | | GG=0 | GG=1 | GG≥2 | **SBx Total** |
| GG=0 | x' | a | d | g | **S0** |
| GG=1 | y' | b | e | h | **S1** |
| GG≥2 | z' | c | f | i | **S2** |
| **MRI Total** | **M0** | **T0** | **T1** | **T2** | |

GG* definition

| | | | | | |
|---|---|---|---|---|---|
| GG*=0 | T0+M0-y'-z' | GG*=1 | d+e+f+y' | GG*≥2 | g+h+i+z' |

MRI+, TBx- | GG*=0

MRI+, TBx+ | GG*=1

MRI+, TBx+ | GG*≥2

**Strategy matrix MRI-SBx, MRI+TBx/SBx (for adjustment)**

| | MRI- | MRI+TBx | | | |
|---|---|---|---|---|---|
| SBx | | GG=0 | GG=1 | GG≥2 | **SBx Total** |
| GG=0 | x | a | d | g | **S0** |
| GG=1 | y | b | e | h | **S1** |
| GG≥2 | z | c | f | i | **S2** |
| **MRI Total** | **M0** | **T0** | **T1** | **T2** | |

GG* definition

GG=0 GG=1 GG≥2

MRI+, TBx-, SBx- | GG=0

MRI+, TBx+, SBx+ | GG=1

MRI+, TBx+, SBx+ | GG≥2

**A1.3 Meta-analyses: Adjustment for probability of positive MRI results given positive PSA and disease stages under strategy *PSA+MRI+TBx/SBx***

**(1) Pr(MRI+ | PSA+, GG=0, Cochrane) / Pr(MRI+ | PSA+, GG*=0, Cochrane)**

= (a/(a+x)) / (a/(a+x+y+z)) = (a+x+y+z)/(a+x)

**(2) Pr(MRI+ | PSA+, GG=1, Cochrane) / Pr(MRI+ | PSA+, GG*=1, Cochrane)**

= ((b+d+e)/(y+b+d+e)) / ((b+d+e)/(b+d+e)) = (b+d+e)/(y+b+d+e)

**(3) Pr(MRI+ | PSA+, GG=2+, Cochrane) / Pr(MRI+ | PSA+, GG*=2+, Cochrane)**

= ((c+f+g+h+i)/(z+c+f+g+h+i)) / ((c+f+g+h+i)/(c+f+g+h+i)) = (c+f+g+h+i)/(z+c+f+g+h+i)

For (1) (2) (3)

GG: defined by Strategy matrix MRI-SBx, MRI+TBx/SBx (for adjustment)

GG*: defined by Strategy matrix PSA+MRI+TBx/SBx

**A1.4 Meta-analyses: Adjustment for probability of positive MRI results given positive PSA and disease stages under strategy *PSA+MRI+TBx***

**(4) Pr(MRI+ | PSA+, GG=0, Cochrane) / Pr(MRI+ | PSA+, GG*=0, Cochrane)**

= (a/(a+x)) / ((a+b+c)/(a+b+c+x+y+z))

**(5) Pr(MRI+ | PSA+, GG=1, Cochrane) / Pr(MRI+ | PSA+, GG*=1, Cochrane)**

= ((b+d+e)/(y+b+d+e)) / ((d+e+f)/(d+e+f)) = (b+d+e)/(y+b+d+e)

**(6) Pr(MRI+ | PSA+, GG=2+, Cochrane) / Pr(MRI+ | PSA+, GG*=2+, Cochrane)**

= ((c+f+g+h+i)/(z+c+f+g+h+i)) / ((g+h+i)/(g+h+i)) = (c+f+g+h+i)/(z+c+f+g+h+i)

For (4) (5) (6)

GG: defined by Strategy matrix MRI-SBx, MRI+TBx/SBx (for adjustment)

GG*: defined by Strategy matrix PSA+MRI+TBx

**A1.5 Adjustment for probability of positive MRI results given positive S3M and disease stages under strategy *S3M+MRI+TBx/SBx***

**(7) Pr(MRI+ | S3M+, GG=0, Phase I) / Pr(MRI+ | S3M+, GG*=0, Phase I)**

= (a/(a+x)) / (a/(a+M0-y'-z')) = (a+M0-y'-z')/(a+x)

**(8) Pr(MRI+ | S3M+, GG=1, Phase I) / Pr(MRI+ | S3M+, GG*=1, Phase I)**

= ((b+d+e)/(y+b+d+e)) / ((b+d+e)/(b+d+e+y')) = (b+d+e+y')/(y+b+d+e)

**(9) Pr(MRI+ | S3M+, GG=1, Phase I) / Pr(MRI+ | S3M+, GG*≥2, Phase I)**

= ((c+f+g+h+i)/(z+c+f+g+h+i)) / ((c+f+g+h+i)/(c+f+g+h+i+z')) = (c+f+g+h+i+z')/(z+c+f+g+h+i)

GG: defined by Strategy matrix MRI-SBx, MRI+TBx/SBx (for adjustment)

GG*: defined by Strategy matrix S3M+MRI+TBx/SBx

**A1.6 Adjustment for probability of positive MRI results given positive S3M and disease stages under strategy *S3M+MRI+TBx***

**(10) Pr(MRI+ | S3M+, GG=0, Phase I) / Pr(MRI+ | S3M+, GG*=0, Phase I)**

= (a/(a+x)) / ((a+b+c)/(a+b+c+M0-y'-z'))

**(11) Pr(MRI+ | S3M+, GG=1, Phase I) / Pr(MRI+ | S3M+, GG*=1, Phase I)**

= ((b+d+e)/(y+b+d+e)) / ((d+e+f)/(d+e+f+y'))

**(12) Pr(MRI+ | S3M+, GG=1, Phase I) / Pr(MRI+ | S3M+, GG*≥2, Phase I)**

= ((c+f+g+h+i)/(z+c+f+g+h+i)) / ((g+h+i)/(g+h+i+z'))

GG: defined by Strategy matrix MRI-SBx, MRI+TBx/SBx (for adjustment)

GG*: defined by Strategy matrix S3M+MRI+TBx

### A2 Results of adjustment parameters

#### A2.1 Meta-analyses: Adjustment for probability of positive MRI results given positive PSA and disease stages under strategy *PSA+MRI+TBx/SBx*

**(1) Pr(MRI+ | PSA+, GG=0, Cochrane) / Pr(MRI+ | PSA+, GG*=0, Cochrane)**

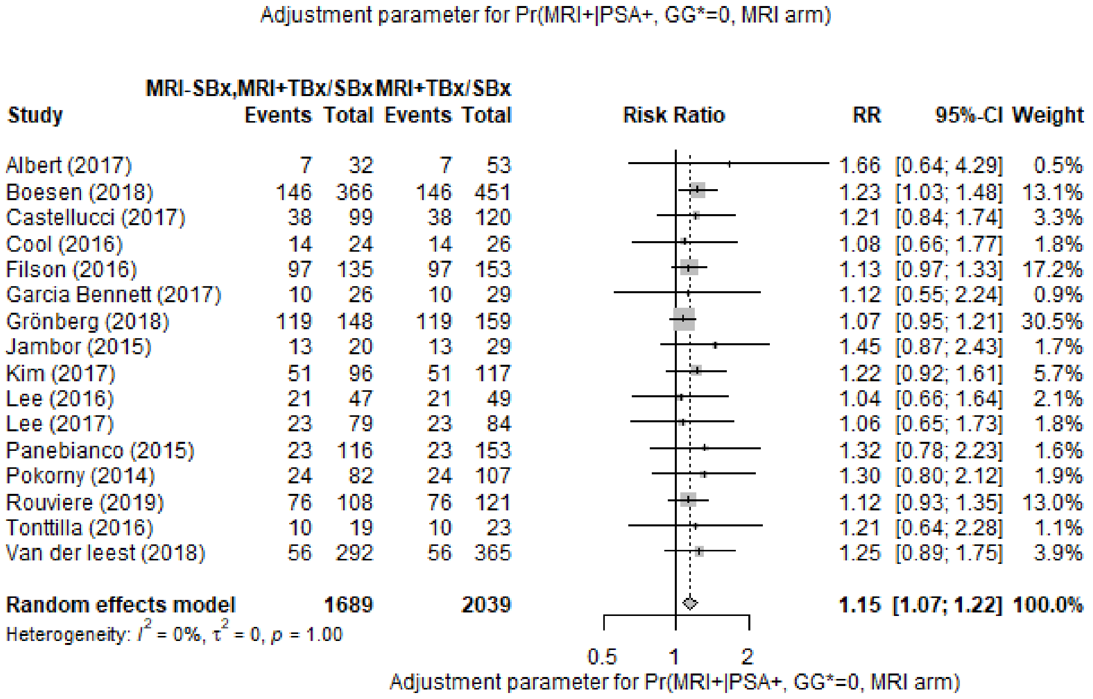

**(2) Pr(MRI+ | PSA+, GG=1, Cochrane) / Pr(MRI+ | PSA+, GG*=1, Cochrane)**

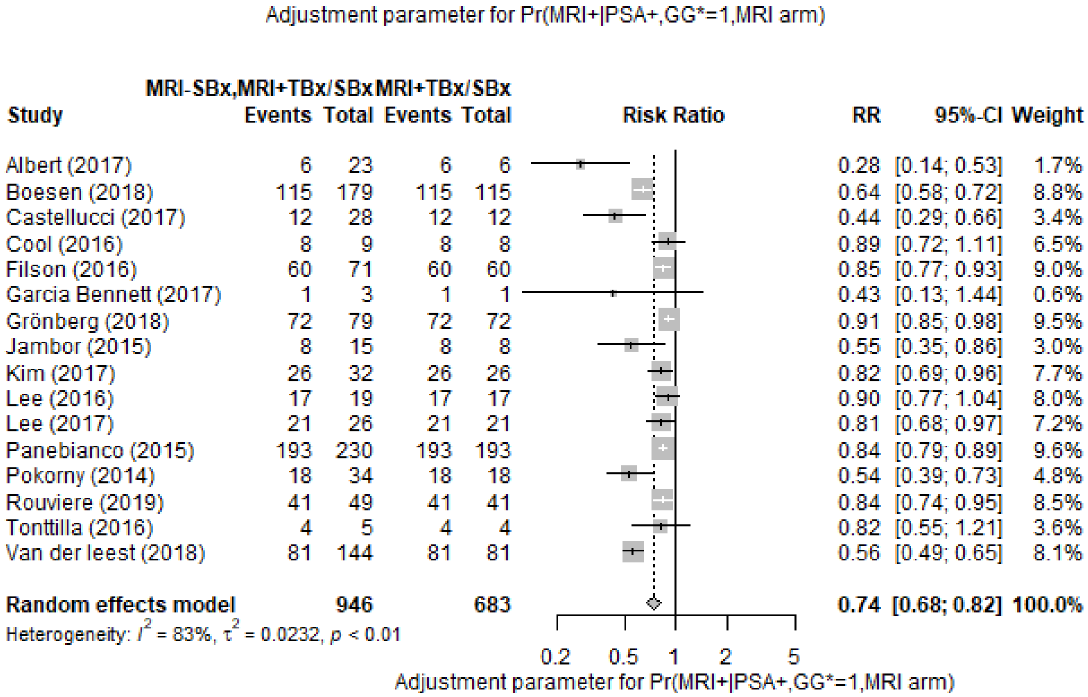

**(3) Pr(MRI+ | PSA+, GG=2+, Cochrane) / Pr(MRI+ | PSA+, GG*=2+, Cochrane)**

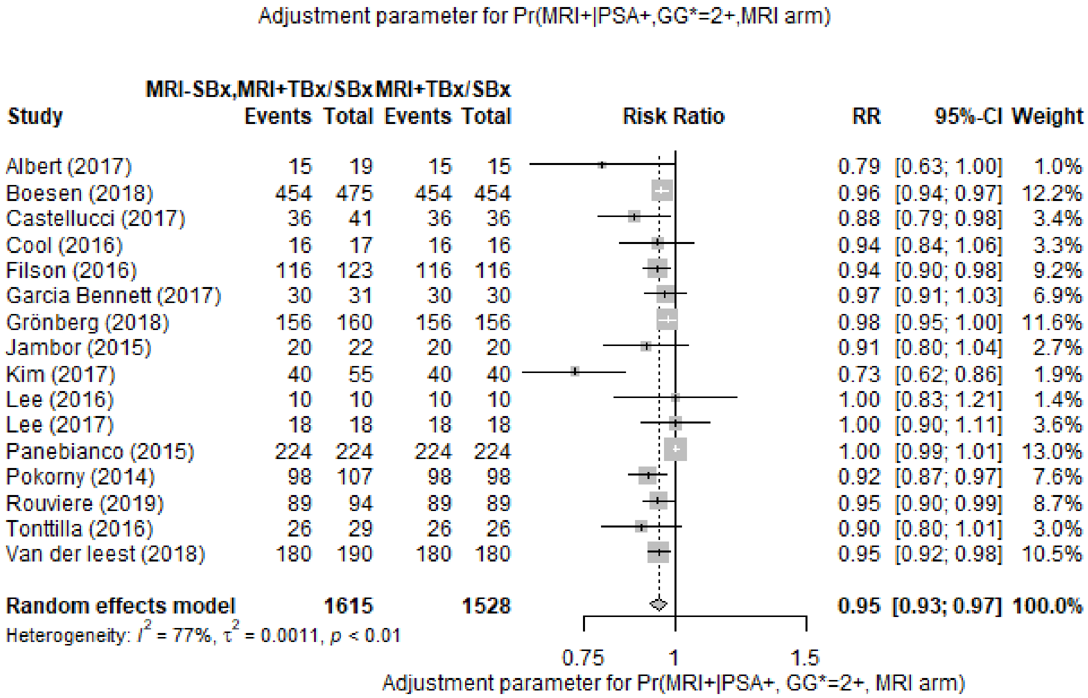

**A2.2 Meta-analyses: Adjustment for probability of positive MRI results given positive PSA and disease stages under strategy *PSA+MRI+TBx***

**(4) Pr(MRI+ | PSA+, GG=0, Cochrane) / Pr(MRI+ | PSA+, GG*=0, Cochrane)**

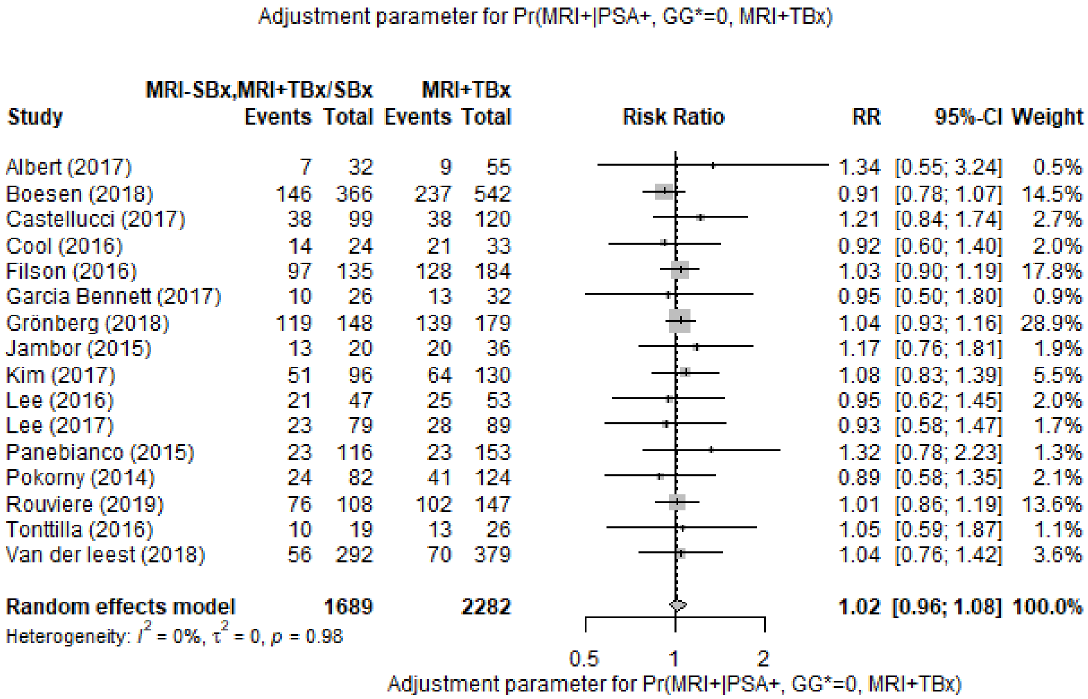

**(5) Pr(MRI+ | PSA+, GG=1, Cochrane) / Pr(MRI+ | PSA+, GG*=1, Cochrane)**

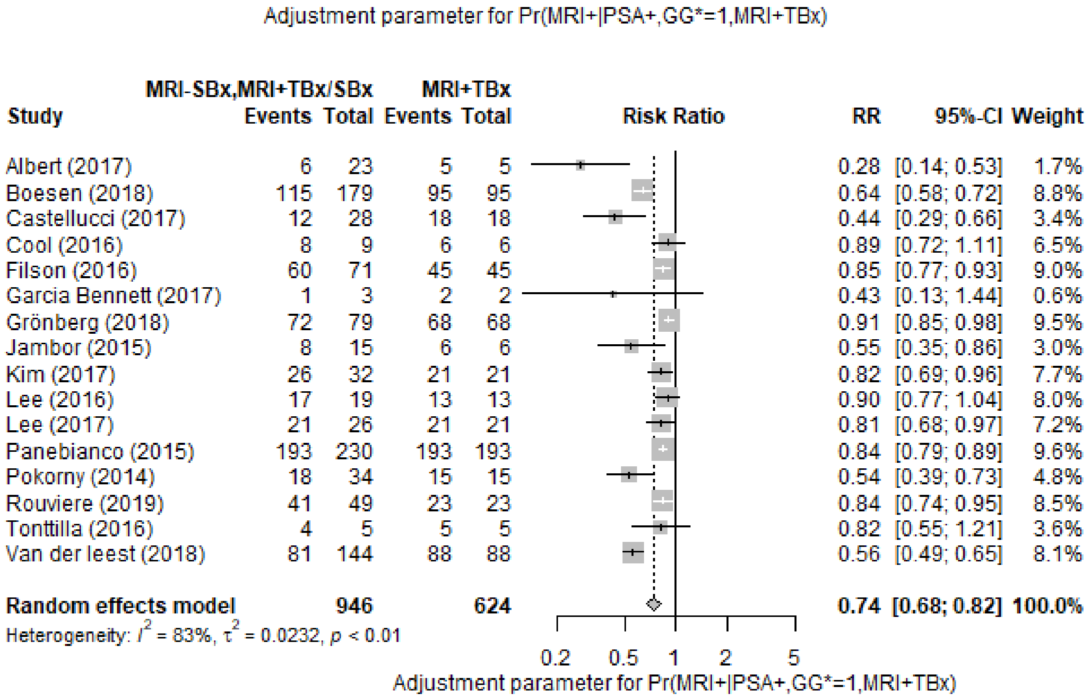

**(6) Pr(MRI+ | PSA+, GG=2+, Cochrane) / Pr(MRI+ | PSA+, GG*=2+, Cochrane)**

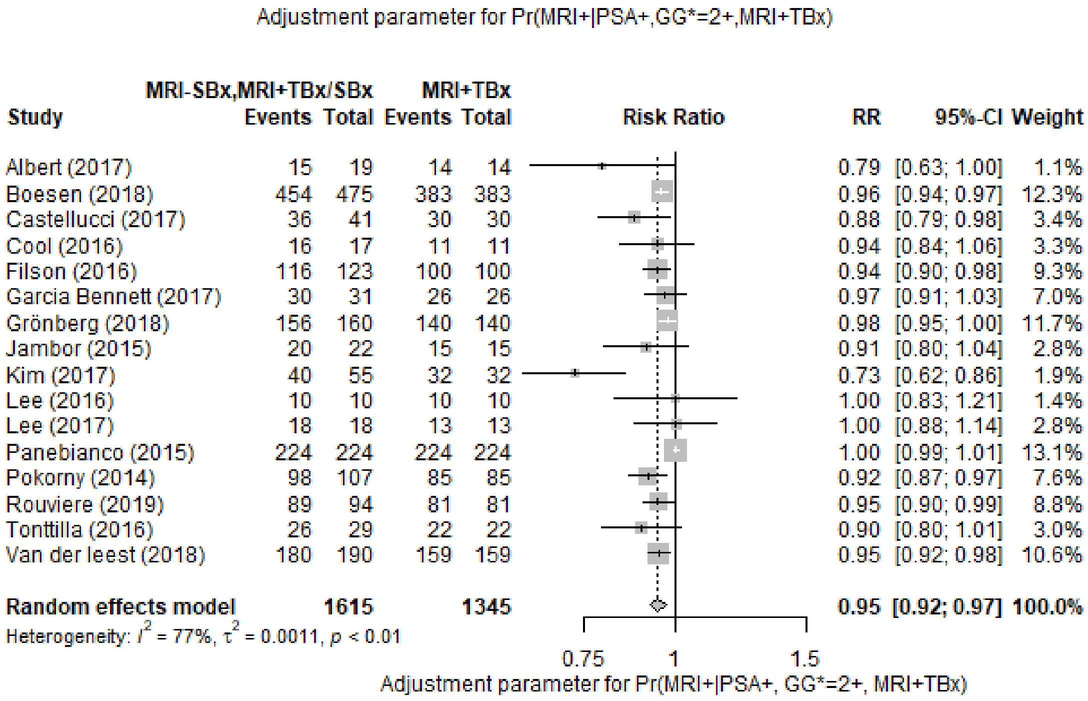

Table S2 Relative positive fractions: S3M+MRI+TBx/SBx using different reflex and S3M test thresholds vs. PSA+MRI+TBx/SBx, MRI-S3M≥25% (per protocol, conditioning on MRI results)

| | ISUP GG=0<br>(PSA≥3 and MRI+) or (PSA≥3 and MRI- and S3M≥25%) | NOT () | ISUP GG=1<br>(PSA≥3 and MRI+) or (PSA≥3 and MRI- and S3M≥25%) | NOT () | ISUP GG≥2<br>(PSA≥3 and MRI+) or (PSA≥3 and MRI- and S3M≥25%) | NOT () |
|---|---|---|---|---|---|---|
| PSA≥3+ and MRI+ | 77 | 0 | 35 | 0 | 177 | 0 |
| NOT () | 23 | 0 | 5 | 0 | 6 | 0 |
| RPF | 1.299 | | 1.143 | | 1.034 | |
| 95% CI | (1.167, 1.446) | | (1.017, 1.285) | | (1.007, 1.062) | |
| p-value | 0.000 | | 0.074 | | 0.041 | |
| | **ISUP GG=0**<br>(PSA≥1.5 and S3M≥15% and MRI+) or (PSA≥1.5 and MRI- and S3M≥25%) | NOT () | **ISUP GG=1**<br>(PSA≥1.5 and S3M≥15% and MRI+) or (PSA≥1.5 and MRI- and S3M≥25%) | NOT () | **ISUP GG≥2**<br>(PSA≥1.5 and S3M≥15% and MRI+) or (PSA≥1.5 and MRI- and S3M≥25%) | NOT () |
| PSA≥3+ and MRI+ | 27 | 50 | 16 | 19 | 145 | 32 |
| NOT () | 53 | 17 | 17 | 7 | 39 | 22 |
| RPF | 1.039 | | 0.943 | | 1.04 | |
| 95% CI | (0.806, 1.339) | | (0.667, 1.333) | | (0.949, 1.139) | |
| p-value | 0.844 | | 0.868 | | 0.476 | |
| | **ISUP GG=0**<br>(PSA≥2 and S3M≥15% and MRI+) or (PSA≥2 and MRI- and S3M≥25%) | NOT () | **ISUP GG=1**<br>(PSA≥2 and S3M≥15% and MRI+) or (PSA≥2 and MRI- and S3M≥25%) | NOT () | **ISUP GG≥2**<br>(PSA≥2 and S3M≥15% and MRI+) or (PSA≥2 and MRI- and S3M≥25%) | NOT () |
| PSA≥3+ and MRI+ | 27 | 50 | 16 | 19 | 145 | 32 |
| NOT () | 43 | 12 | 14 | 3 | 29 | 13 |
| RPF | 0.909 | | 0.857 | | 0.983 | |
| 95% CI | (0.703, 1.176) | | (0.606, 1.213) | | (0.901, 1.073) | |
| p-value | 0.543 | | 0.486 | | 0.798 | |

CI: Confidence Interval; GG: Grade Group; ISUP: International Society of Urological Pathology; MRI: Magnetic Resonance Imaging; PSA: Prostate-Specific Antigen; RFP: Relative Positive Fraction; SBx: Systematic Biopsy; S3M: Stockholm3 test; TBx: Targeted Biopsy; TBx/SBx: Combined Targeted and Systematic Biopsy

Table S3 Relative positive fractions: S3M+MRI+TBx/SBx using different reflex and S3M test thresholds vs. PSA+MRI+TBx/SBx

| | **ISUP GG=0** | | **ISUP GG=1** | | **ISUP GG≥2** | |
|---|---|---|---|---|---|---|
| | PSA≥1.5, S3M≥15% | PSA≥1.5, S3M<15% | PSA≥1.5, S3M≥15% | PSA≥1.5, S3M<15% | PSA≥1.5, S3M≥15% | PSA≥1.5, S3M<15% |
| PSA≥3 | 27 | 50 | 16 | 19 | 145 | 32 |
| PSA<3 | 19 | 17 | 10 | 7 | 31 | 22 |
| RPF | 0.597 | | 0.743 | | 0.994 | |
| 95% CI | (0.454, 0.785) | | (0.524, 1.054) | | (0.91, 1.086) | |
| p-value | 0.000 | | 0.137 | | 1 | |
| | **ISUP GG=0** | | **ISUP GG=1** | | **ISUP GG≥2** | |
| | PSA≥2, S3M≥15% | PSA≥2, S3M<15% | PSA≥2, S3M≥15% | PSA≥2, S3M<15% | PSA≥2, S3M≥15% | PSA≥2, S3M<15% |
| PSA≥3 | 27 | 50 | 16 | 19 | 145 | 32 |
| PSA<3 | 11 | 12 | 8 | 3 | 22 | 13 |
| RPF | 0.494 | | 0.686 | | 0.944 | |
| 95% CI | (0.372, 0.655) | | (0.483, 0.974) | | (0.868, 1.026) | |
| p-value | 0.000 | | 0.054 | | 0.221 | |
| | **ISUP GG=0** | | **ISUP GG=1** | | **ISUP GG≥2** | |
| | PSA≥2, S3M≥11% | PSA≥2, S3M<11% | PSA≥2, S3M≥11% | PSA≥2, S3M<11% | PSA≥2, S3M≥11% | PSA≥2, S3M<11% |
| PSA≥3 | 47 | 30 | 24 | 11 | 158 | 19 |
| PSA<3 | 33 | 0 | 11 | 0 | 35 | 0 |
| RPF | 0.909 | | 1 | | 1.09 | |
| 95% CI | (0.749, 1.104) | | (0.769, 1.3) | | (1.009, 1.179) | |
| p-value | 0.41 | | 1 | | 0.041 | |
| | **ISUP GG=0** | | **ISUP GG=1** | | **ISUP GG≥2** | |
| | PSA≥2.5, S3M≥11% | PSA≥2.5, S3M<11% | PSA≥2.5, S3M≥11% | PSA≥2.5, S3M<11% | PSA≥2.5, S3M≥11% | PSA≥2.5, S3M<11% |
| PSA≥3 | 47 | 30 | 24 | 11 | 158 | 19 |
| PSA<3 | 11 | 0 | 3 | 0 | 20 | 0 |
| RPF | 0.753 | | 0.771 | | 1.006 | |
| 95% CI | (0.624, 0.909) | | (0.608, 0.979) | | (0.939, 1.077) | |
| p-value | 0.005 | | 0.061 | | 1.000 | |
| | **ISUP GG=0** | | **ISUP GG=1** | | **ISUP GG≥2** | |
| | PSA≥2.5, S3M≥15% | PSA≥2.5, S3M<15% | PSA≥2.5, S3M≥15% | PSA≥2.5, S3M<15% | PSA≥2.5, S3M≥15% | PSA≥2.5, S3M<15% |
| PSA≥3 | 27 | 50 | 16 | 19 | 145 | 32 |
| PSA<3 | 7 | 4 | 2 | 1 | 15 | 5 |
| RPF | 0.442 | | 0.514 | | 0.904 | |
| 95% CI | (0.331, 0.59) | | (0.360, 0.736) | | (0.835, 0.979) | |
| p-value | 0.00 | | 0.000 | | 0.020 | |

CI: Confidence Interval; GG: Grade Group; ISUP: International Society of Urological Pathology; MRI: Magnetic Resonance Imaging; PSA: Prostate-Specific Antigen; RFP: Relative P ositive Fraction; SBx: Systematic Biopsy; S3M: Stockholm3 test; TBx: Targeted Biopsy; TBx/SBx: Combined Targeted and Systematic Biopsy

Table S4 Summary of test characteristics and adjustment parameters for base case and one-way sensitivity analysis

| No. | Test characteristics | Value | 95% CI | Adjustment | Value (after adjustment) | 95% CI (after adjustment) |
|---|---|---|---|---|---|---|
| Base case | Pr(MRI+\|PSA+, GG=0, MRI+TBx/SBx) | 0.129 | (0.103, 0.158) | 1.146 | 0.148 | (0.126, 0.192) |
| Base case | Pr(MRI+\|PSA+, GG=1, MRI+TBx/SBx) | 1 | | 0.743 | 0.743 | (0.676, 0.816) |
| Base case | Pr(MRI+\|PSA+, GG≥2, MRI+TBx/SBx) | 1 | | 0.948 | 0.948 | (0.925, 0.971) |
| Base case | Pr(MRI+\|PSA≥1.5, S3M≥15%, GG=0, MRI+TBx/SBx) | 0.145 | (0.108, 0.188) | 1.152 | 0.167 | (0.124, 0.224) |
| Base case | Pr(MRI+\|PSA≥1.5, S3M≥15%, GG=1, MRI+TBx/SBx) | 1 | | 0.960 | 0.960 | (0.796, 0.999) |
| Base case | Pr(MRI+\|PSA≥1.5, S3M≥15%, GG≥2, MRI+TBx/SBx) | 1 | | 0.960 | 0.960 | (0.900, 0.989) |
| Base case | Pr(MRI+\|PSA≥2, S3M≥15%, GG=0, MRI+TBx/SBx) | 0.142 | (0.103, 0.190) | 1.152 | 0.164 | (0.119, 0.226) |
| Base case | Pr(MRI+\|PSA≥2, S3M≥15%, GG=1, MRI+TBx/SBx) | 1 | | 0.960 | 0.960 | (0.796, 0.999) |
| Base case | Pr(MRI+\|PSA≥2, S3M≥15%, GG≥2, MRI+TBx/SBx) | 1 | | 0.959 | 0.959 | (0.899, 0.989) |
| One-way i): S3M≥11% | Pr(MRI+\|PSA≥2, S3M≥11%, GG=0, MRI+TBx/SBx) | 0.133 | (0.105, 0.165) | 1.122 | 0.149 | (0.117, 0.190) |
| One-way i): S3M≥11% | Pr(MRI+\|PSA≥2, S3M≥11%, GG=1, MRI+TBx/SBx) | 1 | | 0.968 | 0.968 | (0.833, 0.999) |
| One-way i): S3M≥11% | Pr(MRI+\|PSA≥2, S3M≥11%, GG≥2, MRI+TBx/SBx) | 1 | | 0.962 | 0.962 | (0.905, 0.990) |
| One-way ii): PSA≥2.5 | Pr(MRI+\|PSA≥2.5, S3M≥11%, GG=0, MRI+TBx/SBx) | 0.144 | (0.111, 0.182) | 1.122 | 0.161 | (0.124, 0.209) |
| One-way ii): PSA≥2.5 | Pr(MRI+\|PSA≥2.5, S3M≥11%, GG=1, MRI+TBx/SBx) | 1 | | 0.968 | 0.968 | (0.833, 0.999) |
| One-way ii): PSA≥2.5 | Pr(MRI+\|PSA≥2.5, S3M≥11%, GG≥2, MRI+TBx/SBx) | 1 | | 0.961 | 0.961 | (0.904, 0.989) |
| One-way ii): PSA≥2.5 | Pr(MRI+\|PSA≥2.5, S3M≥15%, GG=0, MRI+TBx/SBx) | 0.150 | (0.106, 0.204) | 1.152 | 0.173 | (0.124, 0.242) |
| One-way ii): PSA≥2.5 | Pr(MRI+\|PSA≥2.5, S3M≥15%, GG=1, MRI+TBx/SBx) | 1 | | 0.960 | 0.960 | (0.833, 0.999) |
| One-way ii): PSA≥2.5 | Pr(MRI+\|PSA≥2.5, S3M≥15%, GG≥2, MRI+TBx/SBx) | 1 | | 0.958 | 0.958 | (0.904, 0.989) |
| One-way iii): MRI+TBx | Pr(MRI+\|PSA+, GG=0, MRI+TBx) | 0.172 | (0.143, 0.203) | 1.017 | 0.175 | (0.110, 0.454) |
| One-way iii): MRI+TBx | Pr(MRI+\|PSA+, GG=1, MRI+TBx) | 1 | | 0.743 | 0.743 | (0.676, 0.816) |
| One-way iii): MRI+TBx | Pr(MRI+\|PSA+, GG≥2, MRI+TBx) | 1 | | 0.948 | 0.948 | (0.925, 0.971) |
| One-way iii): MRI+TBx | Pr(MRI+\|PSA≥1.5, S3M≥15%, GG=0, MRI+TBx) | 0.200 | (0.159, 0.247) | 1.118 | 0.224 | (0.110, 0.454) |
| One-way iii): MRI+TBx | Pr(MRI+\|PSA≥1.5, S3M≥15%, GG=1, MRI+TBx) | 1 | | 0.960 | 0.960 | (0.796, 0.999) |
| One-way iii): MRI+TBx | Pr(MRI+\|PSA≥1.5, S3M≥15%, GG≥2, MRI+TBx) | 1 | | 0.960 | 0.960 | (0.899, 0.989) |
| One-way iii): MRI+TBx | Pr(MRI+\|PSA≥2, S3M≥15%, GG=0, MRI+TBx) | 0.199 | (0.155, 0.250) | 1.118 | 0.223 | (0.109, 0.456) |
| One-way iii): MRI+TBx | Pr(MRI+\|PSA≥2, S3M≥15%, GG=1, MRI+TBx) | 1 | | 0.960 | 0.960 | (0.796, 0.999) |
| One-way iii): MRI+TBx | Pr(MRI+\|PSA≥2, S3M≥15%, GG≥2, MRI+TBx) | 1 | | 0.959 | 0.959 | (0.898, 0.989) |
| One-way iv): ITT | Pr(MRI+\|PSA+, GG=0, MRI+TBx/SBx) | 0.131 | (0.105, 0.161) | 1.146 | 0.150 | (0.129, 0.195) |
| One-way iv): ITT | Pr(MRI+\|PSA+, GG=1, MRI+TBx/SBx) | 1 | | 0.743 | 0.743 | (0.676, 0.816) |
| One-way iv): ITT | Pr(MRI+\|PSA+, GG≥2, MRI+TBx/SBx) | 1 | | 0.948 | 0.948 | (0.925, 0.971) |
| One-way iv): ITT | Pr(MRI+\|PSA≥1.5, S3M≥15%, GG=0, MRI+TBx/SBx) | 0.152 | (0.114, 0.196) | 1.152 | 0.175 | (0.131, 0.233) |
| One-way iv): ITT | Pr(MRI+\|PSA≥1.5, S3M≥15%, GG=1, MRI+TBx/SBx) | 1 | | 0.960 | 0.960 | (0.796, 0.999) |
| One-way iv): ITT | Pr(MRI+\|PSA≥1.5, S3M≥15%, GG≥2, MRI+TBx/SBx) | 1 | | 0.960 | 0.960 | (0.899, 0.989) |
| One-way iv): ITT | Pr(MRI+\|PSA≥2, S3M≥15%, GG=0, MRI+TBx/SBx) | 0.151 | (0.110, 0.199) | 1.152 | 0.174 | (0.127, 0.236) |
| One-way iv): ITT | Pr(MRI+\|PSA≥2, S3M≥15%, GG=1, MRI+TBx/SBx) | 1 | | 0.960 | 0.960 | (0.796, 0.999) |
| One-way iv): ITT | Pr(MRI+\|PSA≥2, S3M≥15%, GG≥2, MRI+TBx/SBx) | 1 | | 0.959 | 0.959 | (0.898, 0.989) |

CI: Confidence Interval; GG: Grade Group; ISUP: International Society of Urological Pathology; ITT: Intention-to-Treat; MRI: Magnetic Resonance Imaging; PSA: Prostate-Specific Antigen; Pr: Probability; SBx: Systematic Biopsy; S3M: Stockholm3 test; TBx: Targeted Biopsy; TBx/SBx: Combined Targeted and Systematic Biopsy

### Appendix B. Resource and costs characterization

#### B1 Methods

The input data for resources use and unit costs of used resources were extracted from multiple price lists of case in Stockholm Region, a report from the National Board of Health and Welfare (NBHW) and our previous study [5-14]. The duration of the lost production during to diagnosis, treatment, post treatment recovery and terminal care were collected from Heijnsdijk et al [15]. For long-term sick leave by patients with metastatic prostate cancer or patients under palliative therapy, the proportion under employment was 7.68% and the average days of sick leave was approximately 68 days in 2016 [14]. The employment ratio for the general population in Sweden were 0.89, 0.779, 0.175 and 0 for age groups 45-54, 55-64, 65-74 and over 74 years, respectively in 2019 [16]. The average monthly salary of general population in Sweden was €3,211 in 2019 [17], with an additional 37.00% for employer and social contributions [18]. All costs in Swedish kroner were first aligned to the calendar year 2019 by using the consumer price index[19] and then converted to Euro 2019 price by the exchange rate (1 Euro = 10.5892 SEK) from the national bank [20] .

**B2 Results**

Table S5 Resources and unit costs characterization (All costs are at 2019 price)

| Module/Procedure | Unit cost (€) | Resource use | Costs (€) | Source | Lost production | Unit | Years of lost production | Source |
|---|---|---|---|---|---|---|---|---|
| **Diagnosis** | | | | | | | | |
| PSA test at primary care | | | | | | | | |
| GP visit | 144 | 0.20 | 29 | [5-9] | 2 | Hour | 2/24/365.25 | [15] |
| PSA test analysis | 6 | 1 | 6 | [12] | | | | |
| **Total costs** | | | **34** | | **2** | **Hour** | **2/24/365.25** | |
| S3M test at primary care | | | | | | | | |
| GP visit | 144 | 0.2 | 29 | [5-9] | 2 | Hour | 2/24/365.25 | [15] |
| PSA test analysis | 6 | 1 | 6 | [12] | | | | |
| S3M test analysis | 217 | 1 | 217 | A23 lab | | | | |
| **Total costs** | | | **251** | | **2** | **Hour** | **2/24/365.25** | |
| Biopsy at outpatient care: with MRI+TBx | | | | | | | | |
| Specialist and nurse consultation | 140 | 1 | 140 | [12] | 2 | Hour | 2/24/365.25 | [15] |
| Targeted biopsy | 287 | 1 | 287 | [11] | | | | |
| MRI | 336 | 1 | 336 | [12] | 2 | Hour | 2/24/365.25 | Expert opinion |
| Pathology | 407 | 1 | 407 | [12] | | - | - | |
| Nurse consultation | 38 | 1 | 38 | [12] | - | - | - | |
| **Total costs** | | | **1 210** | | **4** | **Hour** | **4/24/365.25** | |
| Biopsy at outpatient care: with MRI+TBx/SBx | | | | | | | | |
| Specialist and nurse consultation | 140 | 1 | 140 | [12] | 2 | Hour | 2/24/365.25 | [15] |
| Systematic and targeted biopsy | 434 | 1 | 434 | [11] | | | | |
| MRI | 336 | 1 | 336 | [12] | 2 | Hour | 2/24/365.25 | Expert opinion |
| Pathology | 407 | 1 | 407 | [12] | | - | - | |
| Nurse consultation | 38 | 1 | 38 | [12] | - | - | - | |
| **Total costs** | | | **1 357** | | **4** | **Hour** | **4/24/365.25** | |
| **Treatment** | | | | | | | | |
| Active surveillance: with MRI+TBx | | | | | | | | |
| Specialist and nurse consultation | 140 | 1 | 140 | | 2 | Hour | 2/24/365.25 | [15] |
| PSA test sampling | 34 | 3 | 103 | [9] | 2*3 | Hour | 2*3/24/365.25 | [15] |
| PSA test analysis | 6 | 3 | 17 | [12] | | | | |
| MRI | 336 | 0.33 | 111 | [12] | 2*0.33 | Hour | 2*0.33/24/365.25 | Expert opinion |

| | | | | | | | | |
|---|---|---|---|---|---|---|---|---|
| Targeted biopsy | 287 | 0.33 | 95 | 11 | 2*0.33 | Hour | 2*0.33/24/365.25 | 15 |
| Pathology | 407 | 0.33 | 134 | 12 | | - | - | |
| **Total cost *(Annual cost)*** | | | **600** | | 9.32 | Hour | **9.32/24/365.25** | |
| Active surveillance: with MRI+TBx/SBx | | | | | | | | |
| Specialist and nurse consultation | 140 | 1 | 140 | | 2 | Hour | 2/24/365.25 | 15 |
| PSA test sampling | 34 | 3 | 103 | 9 | 2*3 | Hour | 2*3/24/365.25 | 15 |
| PSA test analysis | 6 | 3 | 17 | 12 | | | | |
| MRI | 336 | 0.33 | 111 | 12 | 2*0.33 | Hour | 2*0.33/24/365.25 | Expert opinion |
| Systematic and targeted biopsy | 434 | 0.33 | 143 | 11 | 2*0.33 | Hour | 2*0.33/24/365.25 | 15 |
| Pathology | 407 | 0.33 | 134 | 12 | | - | - | |
| **Total cost *(Annual cost)*** | | | **648** | | 9.32 | Hour | **9.32/24/365.25** | |
| Radical prostatectomy: robot-assistant | | | | | | | | |
| Robot assisted surgery | 11 089 | 1 | 11 089 | 13 | | | | |
| Specialist and nurse consultation | 140 | 1 | 140 | 12 | 6 | Week | 6/52 | 15 |
| RT | 12 115 | 0.25 | 3 029 | 13 | | | | |
| **Total cost** | | | **14 258** | | **6** | **Week** | **6/52** | |
| Radiation therapy | | | | | | | | |
| Oncologist consultation - new visit | 375 | 1 | 375 | 8 | | | | |
| Oncologist consultation - further visit | 162 | 1 | 162 | 8 | | | | |
| Nurse visit | 38 | 20 | 769 | 12 | 8 | Week | 8/52 | 15 |
| RT | 606 | 20 | 12 115 | 13 | | | | |
| Hormone therapy-yearly | 6 487 | 0.20 | 1 297 | 14 | | | | |
| **Total cost** | | | **14 719** | | **8** | **Week** | **8/52** | |
| Metastatic: Hormone + chemo therapy | | | | | | | | |
| Year drug cost/patient | 6 880 | 1 | 6 880 | 14 | 67.52 | Day | 67.52/365.25 | 14 |
| **Total cost *(Annual cost)*** | | | **6 880** | | **67.52** | **Day** | **67.52/365.25** | |
| Metastatic: Hormone therapy | | | | | | | | |
| Year drug cost/patient | 6 487 | 1 | 6 487 | 14 | 67.52 | Day | 67.52/365.25 | 14 |
| **Total cost *(Annual cost)*** | | | **6 487** | | **67.52** | **Day** | **67.52/365.25** | |
| **Post treatment follow-up** | | | | | | | | |
| Post treatment follow-up: first year | | | | | | | | |
| Specialist and nurse consultation - Physical | 140 | 1 | 140 | 12 | | | | |
| PSA test sampling | 34 | 1 | 354 | 9 | 2 | Hour | 2/24/365.25 | 15 |
| PSA test analysis | 6 | 1 | 6 | 12 | | | | |
| **Total cost *(Annual cost)*** | | | **180** | | **2** | **Hour** | **2/24/365.25** | |

| | | | | | | | | |
|---|---|---|---|---|---|---|---|---|
| Post treatment follow-up: years following | | | | | | | | |
| Specialist consultation - Tele follow-up | 14 | 1 | 14 | [12] | | | | |
| PSA test sampling | 34 | 1 | 34 | [9] | 2 | Hour | 2/24/365.25 | [15] |
| PSA test analysis | 6 | 1 | 6 | [12] | | | | |
| **Total cost *(Annual cost)*** | | | **54** | | **2** | **Hour** | **2/24/365.25** | |
| **Palliative therapy** | | | | | | | | |
| Yearly drug cost/patient | 15 532 | 1 | 15 532 | [14] | 67.52 | Day | 67.52/365.25 | [14] |
| **Total cost *(Annual cost)*** | | | **15 532** | | **67.52** | **Day** | **67.52/365.25** | |
| **Terminal illness** | | | | | | | | |
| Yearly drug cost/patient | 7 766 | 1 | 7 766 | [14] | 6 | Month | 6/12 | [15] |
| **Total cost** | | | **7 766** | | **6** | **Month** | **6/12** | |

GP: General Practitioner; MRI: Magnetic Resonance Imaging; PSA: Prostate-specific Antigen; RT: Radiation Therapy; SBx: Systematic Biopsy: S3M: Stockholm3 test; TBx: Targeted Biopsy; TBx/SBx: Combined Targeted and Systematic Biopsy

**Appendix C Results of healthcare perspective for base case analysis and results of sensitivity analysis**

Table S6 Summarised predictions in outcomes, costs and ICERs – Base case, healthcare perspective, 3% discounted

| **Part A. Lifetime predictions in outcomes and costs for all strategies** | | | |
|---|---|---|---|
| **Lifetime predictions** | **Strategy I No screening** | **Strategy II PSA3 +MRI+TBx/SBx** | **Strategy IV PSA2+ S3M11%+MRI+TBx** |
| **Outcomes per 10 000 men** | | | |
| MRI | 0 | 33 789 | 33 590 |
| Biopsy | 31 046 | 34 070 | 33 999 |
| Diagnosed PCa | 16 542 | 17 226 | 17 312 |
| Diagnosed PCa – GG≥2 | 11 907 | 12 201 | 12 205 |
| Diagnosed PCa (age 55-69) | 2 708 | 5 126 | 5 354 |
| Diagnosed PCa (age 55-69) – GG≥2 | 1 627 | 2 862 | 2 875 |
| Prostate cancer deaths | 5 774 | 5 280 | 5 239 |
| LY, undiscounted | 2 733 270 | 2 738 022 | 2 738 345 |
| QALYs, undiscounted | 2 203 907 | 2 206 800 | 2 206 979 |
| QALYS, discounted at 3% | 1 476 305 | 1 477 287 | 1 477 331 |
| QALYS, discounted at 5% | 1 182 630 | 1 183 048 | 1 183 057 |
| **Costs (€) per man** | | | |
| Health sector, undiscounted | 362.56 | 395.46 | 396,51 |
| Health sector, discounted at 3% | 177.93 | 211.53 | 213,05 |
| Health sector, discounted at 5% | 116.14 | 148.12 | 149,68 |
| Societal, undiscounted | 371.30 | 411.75 | 413,37 |
| Societal, discounted at 3% | 184.35 | 224.36 | 226,36 |
| Societal, discounted at 5% | 121.46 | 159.22 | 161,21 |

| **Part B. Incremental cost-effectiveness ratios (ICERs) – Healthcare perspective, 3% discounted for QALYs and costs** | | | | |
|---|---|---|---|---|
| **Screening strategy** | **Costs (M€) Per 100,000 men** | **QALYs per 100,000 men** | **ICERs compared to** | |
| | | | **Lowest cost (No screening)** | **Next lowest cost** |
| No screening | 177.93 | 1 476 305 | - | - |
| PSA2+S3M15%+MRI+TBx/SBx | 209.92 | 1 477 260 | 33 479 | 33 479 |
| PSA3+MRI+TBx/SBx | 211.53 | 1 477 287 | 34 233 | 61 864 |
| PSA1.5+S3M15%+MRI+TBx/SBx | 215.69 | 1 477 288 | 38 413 | 2 650 410 |

GG: International Society of Urological Pathology Grade Group; ICER: Incremental Cost-Effectiveness Ratio; LY: Life years; MRI: Magnetic Resonance Imaging; PCa: Prostate Cancer; PSA: Prostate-Specific Antigen; S3M: Stockholm3 test; TBx: Targeted Biopsy; QALY: Quality-Adjusted-Life-Years

Table S7 Summarised predictions in outcomes, costs and ICERs – PSA≥2 ng/mL and S3M≥11%, MRI+TBx/SBx

| **Part A. Lifetime predictions in outcomes and costs for all strategies** | | | |
|---|---|---|---|
| **Lifetime predictions** | **Strategy I No screening** | **Strategy II PSA3 +MRI+TBx/SBx** | **Strategy IV PSA2+ S3M11%+MRI+TBx** |
| **Outcomes per 10 000 men** | | | |
| MRI | 0 | 33 789 | 33 590 |
| Biopsy | 31 046 | 34 070 | 33 999 |
| Diagnosed PCa | 16 542 | 17 226 | 17 312 |
| Diagnosed PCa – GG≥2 | 11 907 | 12 201 | 12 205 |
| Diagnosed PCa (age 55-69) | 2 708 | 5 126 | 5 354 |
| Diagnosed PCa (age 55-69) – GG≥2 | 1 627 | 2 862 | 2 875 |
| Prostate cancer deaths | 5 774 | 5 280 | 5 239 |
| LY, undiscounted | 2 733 270 | 2 738 022 | 2 738 345 |
| QALYs, undiscounted | 2 203 907 | 2 206 800 | 2 206 979 |
| QALYS, discounted at 3% | 1 476 305 | 1 477 287 | 1 477 331 |
| QALYS, discounted at 5% | 1 182 630 | 1 183 048 | 1 183 057 |
| **Costs (€) per man** | | | |
| Health sector, undiscounted | 362.56 | 395.46 | 396,51 |
| Health sector, discounted at 3% | 177.93 | 211.53 | 213,05 |
| Health sector, discounted at 5% | 116.14 | 148.12 | 149,68 |
| Societal, undiscounted | 371.30 | 411.75 | 413,37 |
| Societal, discounted at 3% | 184.35 | 224.36 | 226,36 |
| Societal, discounted at 5% | 121.46 | 159.22 | 161,21 |

| **Part B. Incremental cost-effectiveness ratios (ICERs) – Societal perspective, 3% discounted for QALYs and costs** | | | | |
|---|---|---|---|---|
| **Screening strategy** | **Costs (M€) Per 100,000 men** | **QALYs per 100,000 men** | **ICERs compared to** | |
| | | | **Lowest cost (No screening)** | **Next lowest cost** |
| No screening | 184.35 | 1 476 305 | - | - |
| PSA3+MRI+TBx/SBx | 224.36 | 1 477 287 | 40 764 | 40 764 |
| PSA2+S3M11%+MRI+TBx/SBx | 226.36 | 1 477 331 | 40 941 | 44 823 |

GG: International Society of Urological Pathology Grade Group; ICER: Incremental Cost-Effectiveness Ratio; LY: Life years; MRI: Magnetic Resonance Imaging; PCa: Prostate Cancer; PSA: Prostate-Specific Antigen; S3M: Stockholm3 test; TBx: Targeted Biopsy; QALY: Quality-Adjusted-Life-Years

Table S8 Summarised predictions in outcomes, costs and ICERs – PSA≥2.5ng/mL and S3M≥15% and 11%

| **Lifetime predictions in outcomes and costs for all strategies** | | | | |
|---|---|---|---|---|
| **Lifetime predictions** | **Strategy I No screening** | **Strategy II PSA3 +MRI+TBx/SBx** | **Strategy III PSA2.5+S3M15% +MRI+TBx/SBx** | **Strategy IV PSA2.5+S3M11% +MRI+TBx/SBx** |
| **Outcomes per 100,000 men** | | | | |
| MRI | 0 | 33 789 | 11 748 | 21 681 |
| Biopsy | 31 046 | 34 070 | 31 117 | 32 481 |
| Diagnosed PCa | 16 542 | 17 226 | 16 879 | 17 033 |
| Diagnosed PCa – GG≥2 | 11 907 | 12 201 | 12 136 | 12 197 |
| Diagnosed PCa (age 55-69) | 2 708 | 5 126 | 4 342 | 4 776 |
| Diagnosed PCa (age 55-69) – GG≥2 | 1 627 | 2 862 | 2 714 | 2 858 |
| Prostate cancer deaths | 5 774 | 5 280 | 5 417 | 5 332 |
| LY, undiscounted | 2 733 270 | 2 738 022 | 2 737 024 | 2 737 677 |
| QALYs, undiscounted | 2 203 907 | 2 206 800 | 2 206 310 | 2 206 673 |
| QALYS, discounted at 3% | 1 476 305 | 1 477 287 | 1 477 203 | 1 477 293 |
| QALYS, discounted at 5% | 1 182 630 | 1 183 048 | 1 183 068 | 1 183 084 |
| **Costs (M€) per 100,000 man** | | | | |
| Health sector, undiscounted | 362.56 | 395.46 | 390.83 | 397.59 |
| Health sector, discounted at 3% | 177.93 | 211.53 | 205.59 | 212.31 |
| Health sector, discounted at 5% | 116.14 | 148.12 | 142.19 | 148.48 |
| Societal, undiscounted | 371.30 | 411.75 | 405.04 | 412.94 |
| Societal, discounted at 3% | 184.35 | 224.36 | 216.69 | 224.35 |
| Societal, discounted at 5% | 121.46 | 159.22 | 151.74 | 158.88 |

| **Incremental cost-effectiveness ratios (ICERs) – Societal perspective, 3% discounted for QALYs and costs** | | | | |
|---|---|---|---|---|
| **Strategy** | **Costs (M€) per 100,000 men** | **QALYs per 100,000 men** | **ICERs compared to** | |
| | | | **Lowest cost (No screening)** | **Next lowest cost** |
| No screening | 184.35 | 1 476 305 | - | - |
| PSA2.5+S3M15%+MRI+TBx/SBx | 216.69 | 1 477 203 | 35 997 | 35 997 |
| PSA2.5+S3M11%+MRI+TBx/SBx | 224.35 | 1 477 293 | 40 503 | 85 803 |
| PSA3+MRI+TBx/SBx | 224.36 | 1 477 287 | 40 764 | SD |

GG: International Society of Urological Pathology Grade Group; ICER: Incremental Cost-Effectiveness Ratio; LY: Life years; MRI: Magnetic Resonance Imaging; PCa: Prostate Cancer; PSA: Prostate-Specific Antigen; SBx: Systematic Biopsy; SD: Strongly Dominated; S3M: Stockholm3 test; TBx: Targeted Biopsy; QALY: Quality-Adjusted-Life-Years

Table S9 Summarised predictions in outcomes, costs and ICERs – MRI+TBx

| **Part A. Lifetime predictions in outcomes and costs for all strategies** | | | | |
|---|---|---|---|---|
| **Lifetime predictions** | **Strategy I No screening** | **Strategy II PSA3 +MRI+TBx** | **Strategy III PSA1.5+ S3M15%+MRI+TBx** | **Strategy IV PSA2+ S3M15%+MRI+TBx** |
| **Outcomes per 10 000 men** | | | | |
| MRI | 0 | 34 163 | 13 447 | 11 290 |
| Biopsy | 31 046 | 35 000 | 31 540 | 31 059 |
| Diagnosed PCa | 16 542 | 17 229 | 17 203 | 17 132 |
| Diagnosed PCa – GG≥2 | 11 907 | 12 203 | 12 175 | 12 153 |
| Diagnosed PCa (age 55-69) | 2 708 | 5 135 | 5 150 | 5 004 |
| Diagnosed PCa (age 55-69) – GG≥2 | 1 627 | 2 868 | 2 810 | 2 758 |
| Prostate cancer deaths | 5 774 | 5 279 | 5 268 | 5 293 |
| LY, undiscounted | 2 733 270 | 2 738 033 | 2 738 142 | 2 737 957 |
| QALYs, undiscounted | 2 203 907 | 2 206 801 | 2 206 913 | 2 206 824 |
| QALYS, discounted at 3% | 1 476 305 | 1 477 285 | 1 477 340 | 1 477 326 |
| QALYS, discounted at 5% | 1 182 630 | 1 183 046 | 1 183 082 | 1 183 087 |
| **Costs (€) per man** | | | | |
| Health sector, undiscounted | 362.56 | 395.27 | 402.43 | 395.87 |
| Health sector, discounted at 3% | 177.93 | 211.40 | 217.42 | 211.68 |
| Health sector, discounted at 5% | 116.14 | 148.01 | 153.41 | 148.20 |
| Societal, undiscounted | 371.30 | 411.58 | 418.65 | 411.70 |
| Societal, discounted at 3% | 184.35 | 224.24 | 230.19 | 224.11 |
| Societal, discounted at 5% | 121.46 | 159.12 | 164.45 | 158.95 |

| **Part B. Incremental cost-effectiveness ratios (ICERs) – Societal perspective, 3% discounted for QALYs and costs** | | | | |
|---|---|---|---|---|
| **Strategy** | **Costs (M€) per 100,000 men** | **QALYs per 100,000 men** | **ICERs compared to** | |
| | | | **Lowest cost (No screening)** | **Next lowest cost** |
| No screening | 184.35 | 1 476 305 | - | - |
| PSA2+S3M15%+MRI+TBx | 224.11 | 1 477 326 | 38 958 | 38 958 |
| PSA3+MRI+TBx | 224.24 | 1 477 285 | 40 721 | SD |
| PSA1.5+S3M15%+MRI+TBx | 230.19 | 1 477 340 | 44 313 | 108 413 |

GG: International Society of Urological Pathology Grade Group; ICER: Incremental Cost-Effectiveness Ratio; LY: Life years; MRI: Magnetic Resonance Imaging; PCa: Prostate Cancer; PSA: Prostate-Specific Antigen; SBx: Systematic Biopsy; SD: Strongly Dominated; S3M: Stockholm3 test; TBx: Targeted Biopsy; QALY: Quality-Adjusted-Life-Years

Table S10 Summarised predictions in outcomes, costs and ICERs – Test characteristics using ITT method

| **Part A. Lifetime predictions in outcomes and costs for all strategies** | | | | |
|---|---|---|---|---|
| **Lifetime predictions** | **Strategy I No screening** | **Strategy II PSA3 +MRI+TBx/SBx, ITT** | **Strategy III PSA1.5+ S3M15%+MRI+TBx/SBx, ITT** | **Strategy IV PSA2+ S3M15%+MRI+TBx/SBx, ITT** |
| **Outcomes per 10 000 men** | | | | |
| MRI | 0 | 33 818 | 21 631 | 18 782 |
| Biopsy | 31 046 | 34 140 | 32 707 | 32 229 |
| Diagnosed PCa | 16 542 | 17 226 | 17 054 | 16 991 |
| Diagnosed PCa – GG≥2 | 11 907 | 12 201 | 12 193 | 12 160 |
| Diagnosed PCa (age 55-69) | 2 708 | 5 127 | 4 834 | 4 678 |
| Diagnosed PCa (age 55-69) – GG≥2 | 1 627 | 2 863 | 2 852 | 2 777 |
| Prostate cancer deaths | 5 774 | 5 280 | 5 321 | 5 353 |
| LY, undiscounted | 2 733 270 | 2 738 023 | 2 737 766 | 2 737 520 |
| QALYs, undiscounted | 2 203 907 | 2 206 800 | 2 206 724 | 2 206 587 |
| QALYS, discounted at 3% | 1 476 305 | 1 477 286 | 1 477 306 | 1 477 273 |
| QALYS, discounted at 5% | 1 182 630 | 1 183 048 | 1 183 087 | 1 183 082 |
| **Costs (€) per man** | | | | |
| Health sector, undiscounted | 362.56 | 395.54 | 405.34 | 398.68 |
| Health sector, discounted at 3% | 177.93 | 211.59 | 218.86 | 212.90 |
| Health sector, discounted at 5% | 116.14 | 148.18 | 154.36 | 148.92 |
| Societal, undiscounted | 371.30 | 411.83 | 420.81 | 413.72 |
| Societal, discounted at 3% | 184.35 | 224.43 | 231.01 | 224.69 |
| Societal, discounted at 5% | 121.46 | 159.28 | 164.85 | 159.08 |

| **Part B. Incremental cost-effectiveness ratios (ICERs) – Societal perspective, 3% discounted for QALYs and costs** | | | | |
|---|---|---|---|---|
| **Strategy** | **Costs (M€) per 100,000 men** | **QALYs per 100,000 men** | **ICERs compared to** | |
| | | | **Lowest cost (No screening)** | **Next lowest cost** |
| No screening | 184.35 | 1 476 305 | - | - |
| PSA3+MRI+TBx/SBx, ITT | 224.43 | 1 477 286 | 40 839 | 40 839 |
| PSA2+S3M15%+MRI+TBx/SBx, ITT | 224.69 | 1 477 273 | 41 697 | SD |
| PSA1.5+S3M15%+MRI+TBx/SBx, ITT | 231.01 | 1 477 306 | 46 630 | 190 314 |

GG: International Society of Urological Pathology Grade Group; ICER: Incremental Cost-Effectiveness Ratio; ITT: Intention-To-Treat; LY: Life years; MRI: Magnetic Resonance Imaging; PCa: Prostate Cancer; PSA: Prostate-Specific Antigen; SD: Strongly Dominated; SBx: Systematic Biopsy; S3M: Stockholm3 test; TBx: Targeted Biopsy; QALY: Quality-Adjusted-Life-Years

Table S11 Summarised predictions in outcomes, costs and ICERs – Lower unit cost of S3M test (€94)

| **Lifetime predictions in outcomes and costs for all strategies** | | | | |
|---|---|---|---|---|
| **Lifetime predictions** | **Strategy I No screening** | **Strategy II PSA3 +MRI+TBx/SBx** | **Strategy III PSA1.5+S3M15% +MRI+TBx/SBx, lower S3M** | **Strategy IV PSA2+S3M15% +MRI+TBx/SBx, lower S3M** |
| **Outcomes per 100,000 men** | | | | |
| MRI | 0 | 33 789 | 16 226 | 13 439 |
| Biopsy | 31 046 | 34 070 | 31 632 | 31 162 |
| Diagnosed PCa | 16 542 | 17 226 | 17 011 | 16 957 |
| Diagnosed PCa – GG≥2 | 11 907 | 12 201 | 12 186 | 12 156 |
| Diagnosed PCa (age 55-69) | 2 708 | 5 126 | 4 720 | 4 582 |
| Diagnosed PCa (age 55-69) – GG≥2 | 1 627 | 2 862 | 2 835 | 2 764 |
| Prostate cancer deaths | 5 774 | 5 280 | 5 342 | 5 370 |
| LY, undiscounted | 2 733 270 | 2 738 022 | 2 737 605 | 2 737 393 |
| QALYs, undiscounted | 2 203 907 | 2 206 800 | 2 206 640 | 2 206 523 |
| QALYS, discounted at 3% | 1 476 305 | 1 477 287 | 1 477 288 | 1 477 260 |
| QALYS, discounted at 5% | 1 182 630 | 1 183 048 | 1 183 087 | 1 183 083 |
| **Costs (M€) per 100,000 man** | | | | |
| Health sector, undiscounted | 362.56 | 395.46 | 391.70 | 387.71 |
| Health sector, discounted at 3% | 177.93 | 211.53 | 207.65 | 203.97 |
| Health sector, discounted at 5% | 116.14 | 148.12 | 144.48 | 141.10 |
| Societal, undiscounted | 371.30 | 411.75 | 406.88 | 402.51 |
| Societal, discounted at 3% | 184.35 | 224.36 | 219.55 | 215.57 |
| Societal, discounted at 5% | 121.46 | 159.22 | 154.76 | 151.10 |

| **Incremental cost-effectiveness ratios (ICERs) – Societal perspective, 3% discounted for QALYs and costs** | | | | |
|---|---|---|---|---|
| **Strategy** | **Costs (M€) per 100,000 men** | **QALYs per 100,000 men** | **ICERs compared to** | |
| | | | **Lowest cost (No screening)** | **Next lowest cost** |
| No screening | 184.35 | 1 476 305 | - | - |
| PSA2+S3M15%+MRI+TBx/SBx, lower S3M cost | 215.57 | 1 477 260 | 32 671 | 32 671 |
| PSA1.5+S3M15%+MRI+TBx/SBx, lower S3M cost | 219.55 | 1 477 288 | 35 809 | 144 307 |
| PSA3+MRI+TBx/SBx | 224.36 | 1 477 287 | 40 764 | SD |

GG: International Society of Urological Pathology Grade Group; ICER: Incremental Cost-Effectiveness Ratio; ITT: Intention-To-Treat; LY: Life years; MRI: Magnetic Resonance Imaging; PCa: Prostate Cancer; PSA: Prostate-Specific Antigen; SBx: Systematic Biopsy; S3M: Stockholm3 test; TBx: Targeted Biopsy; QALY: Quality-Adjusted-Life-Years

Table S12 Summarised predictions in outcomes, costs and ICERs – Higher unit cost of S3M test (€283)

| **Lifetime predictions in outcomes and costs for all strategies** | | | | |
|---|---|---|---|---|
| **Lifetime predictions** | **Strategy I No screening** | **Strategy II PSA3 +MRI+TBx/SBx** | **Strategy III PSA1.5+S3M15% +MRI+TBx/SBx, higher S3M** | **Strategy IV PSA2+S3M15% +MRI+TBx/SBx, higher S3M** |
| **Outcomes per 100,000 men** | | | | |
| MRI | 0 | 33 789 | 16 226 | 13 439 |
| Biopsy | 31 046 | 34 070 | 31 632 | 31 162 |
| Diagnosed PCa | 16 542 | 17 226 | 17 011 | 16 957 |
| Diagnosed PCa – GG≥2 | 11 907 | 12 201 | 12 186 | 12 156 |
| Diagnosed PCa (age 55-69) | 2 708 | 5 126 | 4 720 | 4 582 |
| Diagnosed PCa (age 55-69) – GG≥2 | 1 627 | 2 862 | 2 835 | 2 764 |
| Prostate cancer deaths | 5 774 | 5 280 | 5 342 | 5 370 |
| LY, undiscounted | 2 733 270 | 2 738 022 | 2 737 605 | 2 737 393 |
| QALYs, undiscounted | 2 203 907 | 2 206 800 | 2 206 640 | 2 206 523 |
| QALYS, discounted at 3% | 1 476 305 | 1 477 287 | 1 477 288 | 1 477 260 |
| QALYS, discounted at 5% | 1 182 630 | 1 183 048 | 1 183 087 | 1 183 083 |
| **Costs (M€) per 100,000 man** | | | | |
| Health sector, undiscounted | 362.56 | 395.46 | 407.13 | 399.25 |
| Health sector, discounted at 3% | 177.93 | 211.53 | 220.02 | 213.12 |
| Health sector, discounted at 5% | 116.14 | 148.12 | 155.30 | 149.03 |
| Societal, undiscounted | 371.30 | 411.75 | 422.31 | 414.06 |
| Societal, discounted at 3% | 184.35 | 224.36 | 231.93 | 224.71 |
| Societal, discounted at 5% | 121.46 | 159.22 | 165.58 | 159.03 |

| **Incremental cost-effectiveness ratios (ICERs) – Societal perspective, 3% discounted for QALYs and costs** | | | | |
|---|---|---|---|---|
| **Strategy** | **Costs (M€) per 100,000 men** | **QALYs per 100,000 men** | **ICERs compared to** | |
| | | | **Lowest cost (No screening)** | **Next lowest cost** |
| No screening | 184,35 | 1 476 305 | - | - |
| PSA3+MRI+TBx/SBx | 224,36 | 1 477 287 | 40 764 | 40 764 |
| PSA2+S3M15%+MRI+TBx/SBx, higher S3M cost | 224,71 | 1 477 260 | 42 245 | SD |
| PSA1.5+S3M15%+MRI+TBx/SBx, higher S3M cost | 231,93 | 1 477 288 | 48 399 | SD |

GG: International Society of Urological Pathology Grade Group; ICER: Incremental Cost-Effectiveness Ratio; ITT: Intention-To-Treat; LY: Life years; MRI: Magnetic Resonance Imaging; PCa: Prostate Cancer; PSA: Prostate-Specific Antigen; SBx: Systematic Biopsy; S3M: Stockholm3 test; TBx: Targeted Biopsy; QALY: Quality-Adjusted-Life-Years

Table S13 Summarised predictions in outcomes, costs and ICERs – Background HSV using value sets for country A from WHO

| Lifetime predictions in outcomes and costs for all strategies | | | | |
|---|---|---|---|---|
| **Lifetime predictions** | **Strategy I No screening** | **Strategy II PSA3 +MRI+TBx/SBx** | **Strategy III PSA1.5+S3M15% +MRI+TBx/SBx, higher S3M** | **Strategy IV PSA2+S3M15% +MRI+TBx/SBx, higher S3M** |
| **Outcomes per 100,000 men** | | | | |
| MRI | 0 | 33 787 | 16 221 | 13 439 |
| Biopsy | 31 046 | 34 072 | 31 633 | 31 166 |
| Diagnosed PCa | 16 542 | 17 226 | 17 011 | 16 957 |
| Diagnosed PCa – GG≥2 | 11 907 | 12 201 | 12 187 | 12 156 |
| Diagnosed PCa (age 55-69) | 2 708 | 5 127 | 4 717 | 4 579 |
| Diagnosed PCa (age 55-69) – GG≥2 | 1 627 | 2 862 | 2 834 | 2 765 |
| Prostate cancer deaths | 5 774 | 5 279 | 5 342 | 5 370 |
| LY, undiscounted | 2 733 270 | 2 738 021 | 2 737 601 | 2 737 389 |
| QALYs, undiscounted | 2 119 287 | 2 121 719 | 2 121 601 | 2 121 506 |
| QALYS, discounted at 3% | 1 447 421 | 1 448 203 | 1 448 221 | 1 448 202 |
| QALYS, discounted at 5% | 1 172 673 | 1 172 973 | 1 173 020 | 1 173 021 |
| **Costs (M€) per 100,000 man** | | | | |
| Health sector. undiscounted | 362.56 | 395.45 | 401.68 | 395.19 |
| Health sector. discounted at 3% | 177.93 | 211.53 | 215.66 | 209.90 |
| Health sector. discounted at 5% | 116.14 | 148.12 | 151.49 | 146.24 |
| Societal. undiscounted | 371.30 | 411.74 | 416.85 | 409.99 |
| Societal. discounted at 3% | 184.35 | 224.36 | 227.56 | 221.49 |
| Societal. discounted at 5% | 121.46 | 159.22 | 161.76 | 156.23 |

| Incremental cost-effectiveness ratios (ICERs) – Societal perspective. 3% discounted for QALYs and costs | | | | |
|---|---|---|---|---|
| **Strategy** | **Costs (M€) per 100.000 men** | **QALYs per 100.000 men** | **ICERs compared to** | |
| | | | **Lowest cost (No screening)** | **Next lowest cost** |
| No screening | 184,35 | 1 447 421 | - | - |
| PSA2+S3M15%+MRI+TBx/SBx, WHO HSV | 221,49 | 1 448 202 | 47 579 | 47 579 |
| PSA3+MRI+TBx/SBx, WHO HSV | 224,36 | 1 448 203 | 51 161 | 2 024 168 |
| PSA1.5+S3M15%+MRI+TBx/SBx, WHO HSV | 227,56 | 1 448 221 | 54 023 | 179 274 |

GG: International Society of Urological Pathology Grade Group; ICER: Incremental Cost-Effectiveness Ratio; ITT: Intention-To-Treat; LY: Life years; MRI: Magnetic Resonance Imaging; PCa: Prostate Cancer; PSA: Prostate-Specific Antigen; SBx: Systematic Biopsy; S3M: Stockholm3 test; TBx: Targeted Biopsy; QALY: Quality-Adjusted-Life-Years

Table S14 Summarised predictions in outcomes. costs and ICERs – Discount rate 0% and 5%

| **Incremental cost-effectiveness ratios (ICERs) – Societal perspective, no discount for QALYs and costs** | | | | | |
|---|---|---|---|---|---|
| **Strategy** | **Costs (M€) per 100,000 men** | **QALYs per 100,000 men** | **ICERs compared to** | | |
| | | | **Lowest cost (No screening)** | **Next lowest cost** | **Relative alternative** |
| No screening | 371.30 | 2 203 907 | - | - | - |
| PSA2+S3M15%+MRI+TBx/SBx | 410.02 | 2 206 523 | 14 799 | 14 799 | ED |
| PSA3+MRI+TBx/SBx | 411.75 | 2 206 800 | 13 983 | 6 262 | 13 983 |
| PSA1.5+S3M15%+MRI+TBx/SBx | 416.91 | 2 206 640 | 16 690 | SD | 16 690 |

ICER: Incremental Cost-Effectiveness Ratio; ITT: Intention-To-Treat; LY: Life years; MRI: Magnetic Resonance Imaging; PCa: Prostate Cancer; PSA: Prostate-Specific Antigen; SBx: Systematic Biopsy; S3M: Stockholm3 test; TBx: Targeted Biopsy; QALY: Quality-Adjusted-Life-Years

| **Incremental cost-effectiveness ratios (ICERs) – Societal perspective, 3% discounted for QALYs and costs** | | | | |
|---|---|---|---|---|
| **Strategy** | **Costs (M€) per 100,000 men** | **QALYs per 100,000 men** | **ICERs compared to** | |
| | | | **Lowest cost (No screening)** | **Next lowest cost** |
| No screening | 121.46 | 1 182 630 | - | - |
| PSA2+S3M15%+MRI+TBx/SBx | 156.25 | 1 183 048 | 83 114 | 83 114 |
| PSA3+MRI+TBx/SBx | 159.22 | 1 183 083 | 83 316 | 85 755 |
| PSA1.5+S3M15%+MRI+TBx/SBx | 161.79 | 1 183 087 | 88 206 | 643 461 |

ICER: Incremental Cost-Effectiveness Ratio; ITT: Intention-To-Treat; LY: Life years; MRI: Magnetic Resonance Imaging; PCa: Prostate Cancer; PSA: Prostate-Specific Antigen; SBx: Systematic Biopsy; S3M: Stockholm3 test; TBx: Targeted Biopsy; QALY: Quality-Adjusted-Life-Years

Figure S2 Cost-effectiveness plane of the probabilistic sensitivity analysis

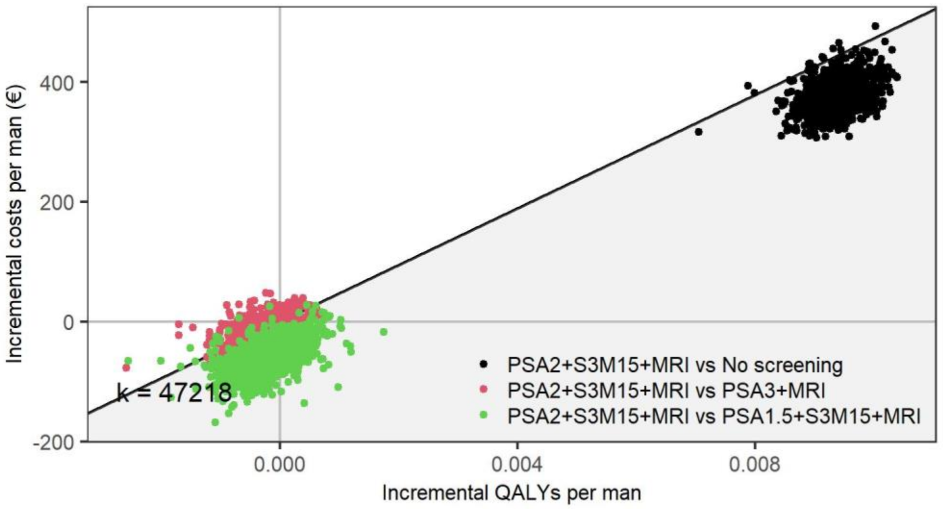

MRI: Magnetic Resonance Imaging; QALYs: Quality-Adjusted Life Years

Figure S3 Expected value of perfect information by cost-effectiveness threshold, societal perspective, 3% discounted

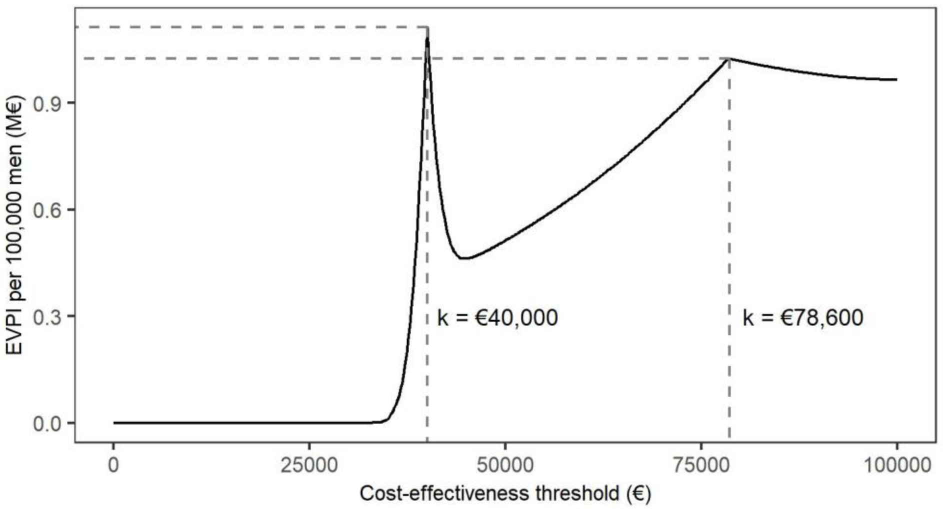

The EVPI firstly peaked at a cost-effectiveness threshold was €40,000 per QALY gained, where the optimal strategy changed from no screening to PSA2+S3M15+MRI on the cost-effectiveness acceptability curve (Figure 4). At a cost-effectiveness threshold of €47,218 per QALY gained, the PSA2+S3M15+MRI strategy was not expected to be cost-effective and the EVPI was close to €0.5M per 100,000 men. If the threshold is greater than €78,600 per QALY gained, the probability that PSA2+S3M15+MRI decreased to 50%. EVPI: Expected Value of Perfect Information; M: Million

### References

1. Drost FH, Osses DF, Nieboer D, et al. Prostate MRI, with or without MRI-targeted biopsy, and systematic biopsy for detecting prostate cancer. *The Cochrane database of systematic reviews.* 2019;4:Cd012663.
2. Nordström T, Jäderling F, Carlsson S, Aly M, Grönberg H, Eklund M. Does a novel diagnostic pathway including blood-based risk prediction and MRI-targeted biopsies outperform prostate cancer screening using prostate-specific antigen and systematic prostate biopsies? - protocol of the randomised study STHLM3MRI. *BMJ Open.* 2019;9(6):e027816.
3. European Association of Urology. *Oncology guidelines prostate cancer.* Arnhem, The Netherlands: European Association of Urology;2019.
4. Hosseiny M, Raman SS. MRI-Guided In-Bore and MRI-Targeted US (Fusion) Biopsy. In: Tirkes T, ed. *Prostate MRI Essentials.* Springer, Cham; 2020:129-145.
5. Socialstyrelsen. Öppna jämförelser 2014 Hälso- och sjukvårdHälso- och sjukvård. Jämförelser mellan landsting. Del 1. Övergripande indikatorer. In:2014.
6. Socialstyrelsen. Öppna jämförelser 2015 Hälso- och sjukvård. Övergripande indikatorer. In:2015.
7. SödraRegionvårdsnämnden. Regionala priser och ersättningar för södra sjukvårdsregionen 2015. In:2015.
8. SödraRegionvårdsnämnden. Regionala priser och ersättningar för södra sjukvårdsregionen 2016. In:2016.
9. Socialstyrelsen. Screening för prostatacancer med PSA-prov. Hälsoekonomisk analys Bilaga. 2018. Published February 13, 2018. Accessed May 21, 2018.
10. SLL. *Avtal med Karolinska sjukhuset avseende tjänster inom klinisk laboratoriemedicin i sydöstra länet HSN 2018-0963.* Stockholm2018.
11. Rapporteringsanvisningar Urologi, specialiserad. 2019. https://vardgivarguiden.se/administration/verksamhetsadministration/rapportera/rapporteringsanvisningar-a-o/urologi-specialiserad/? Accessed 2019-11-29.
12. SLL. *Vårdval specialiserad urologi.* Aug. 1 2018.
13. Socialstyrelsen. *Hälsoekonomiskt underlag. Nationella riktlinjer för prostatacancer 2014.* 2014.
14. Hao S, Östensson E, Eklund M, et al. The economic burden of prostate cancer – a Swedish prevalence-based register study. *BMC Health Services Research.* 2020;20(1):448.
15. Heijnsdijk EA, de Carvalho TM, Auvinen A, et al. Cost-effectiveness of prostate cancer screening: a simulation study based on ERSPC data. *Journal of the National Cancer Institute.* 2015;107(1):366.
16. Population aged 15-74 (LFS) by sex, age and labour status. Year 1970 - 2019. 2020. http://www.statistikdatabasen.scb.se/pxweb/en/ssd/START__AM__AM0401__AM0401A/NAKUBefolkning2Ar/. Accessed 2020-12-02.
17. Salary structures, whole economy. SCB; 2020. http://www.statistikdatabasen.scb.se/pxweb/en/ssd/START__AM__AM0110__AM0110A/LonYrkeUtbildning4A/. Accessed 2020-12-02.
18. Carlgren F. Sociala avgifter över tid - Avtalade och lagstadgade avgifter - arbetare. ekonomifakta. https://www.ekonomifakta.se/Fakta/Skatter/Skatt-pa-arbete/Sociala-avgifter-over-tid/. Published 2020. Updated 2020-06-30. Accessed 2019-12-02.
19. SCB. CPI, Fixed Index Numbers (1980=100). SCB. https://www.scb.se/en/finding-statistics/statistics-by-subject-area/prices-and-consumption/consumer-price-index/consumer-price-index-cpi/pong/tables-and-graphs/consumer-price-index-cpi/cpi-fixed-index-numbers-1980100/. Published 2020. Updated 2020-11-12. Accessed 2020-12-02, 2020.
20. Annual average exchange rates. Swedish National Bank; 2020. https://www.riksbank.se/en-gb/statistics/search-interest--exchange-rates/annual-average-exchange-rates/. Accessed 2020-12-02.
