## Supplementary Material II for "Cost-effectiveness of Stockholm3 test and magnetic resonance imaging in prostate cancer screening: a microsimulation study"

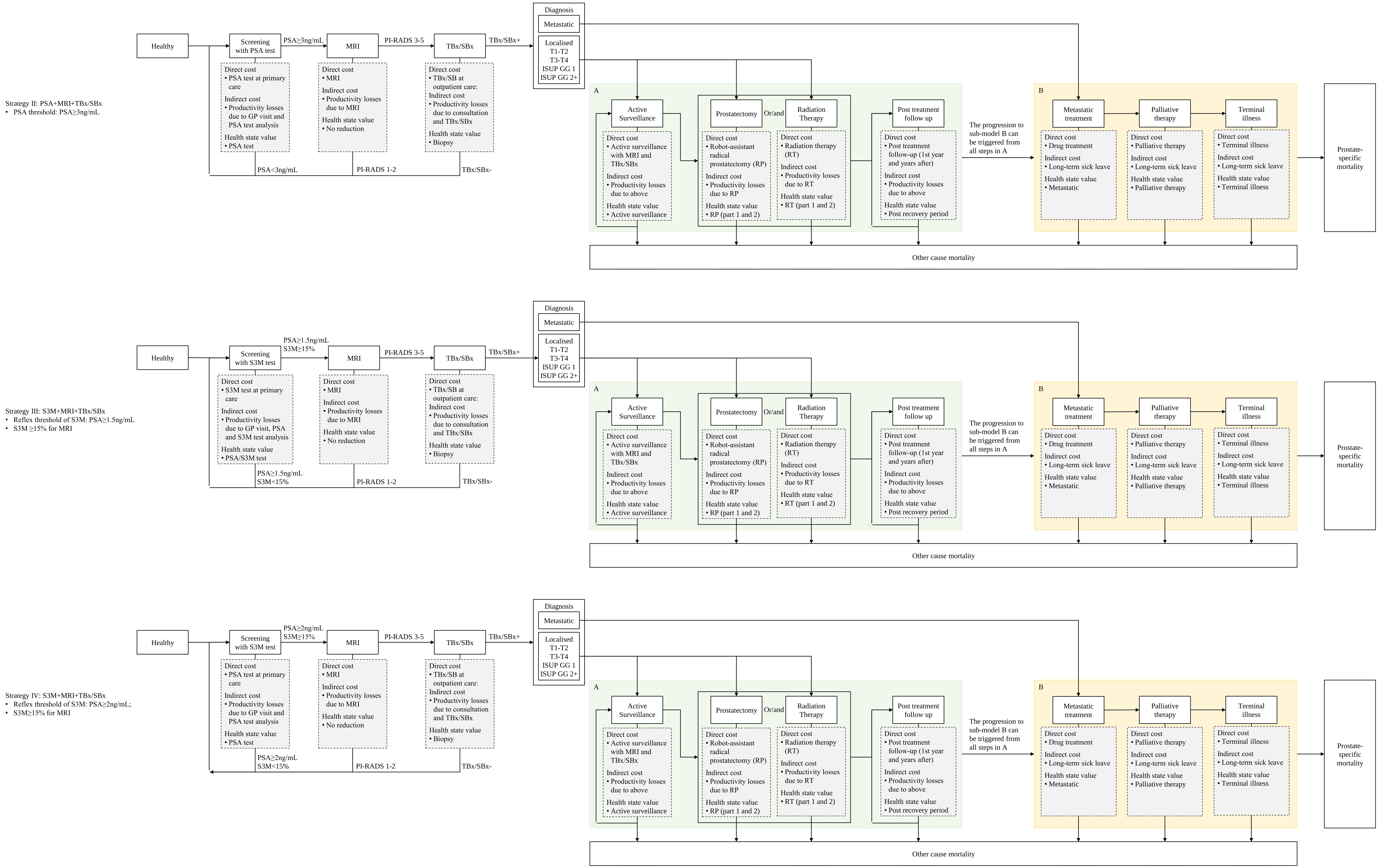


Sub-model A: Treatments for patients with localised prostate cancer
Sub-model B: Treatments for patients with metastatic cancer; Palliative therapy and Terminal care.
GG: ISUP Grade Group; GP: General Practitioner; ISUP: International Society of Urological Pathology; MRI: Magnetic Resonance Imaging; PI-RADS: Prostate Imaging–Reporting and Data System; PSA: Prostate-Specific Antigen; RP: Radical Prostatectomy; RT: Radiation Therapy; S3M: Stockholm 3 test; SBx: Systematic Biopsy; TBx: MRI-guided Targeted Biopsy; TBx/SBx: combined Targeted and Systematic Biopsies
